# Age-related clonal haematopoiesis is more prevalent in older adults with HIV: the ARCHIVE study

**DOI:** 10.1101/2020.11.19.20235069

**Authors:** Nila J. Dharan, Paul Yeh, Mark Bloch, Miriam Yeung, David Baker, Jerick Guinto, Norm Roth, Sarah Ftouni, Katherine Ognenovska, Don Smith, Jennifer F. Hoy, Ian Woolley, Catherine Pell, David J. Templeton, Neil Fraser, Nectarios Rose, Jolie Hutchinson, Kathy Petoumenos, Sarah-Jane Dawson, Mark N. Polizzotto, Mark A. Dawson, for the ARCHIVE Study Group

## Abstract

People with HIV have higher rates of certain comorbidities, particularly cardiovascular disease and some malignancies, than people without HIV. As somatic mutations associated with age-related clonal haematopoiesis (CH) are linked to similar comorbidities in the general population, we hypothesized that CH may be more prevalent in people with HIV. To address this issue, we established a prospective cohort study recruiting 220 HIV-positive and 226 HIV-negative participants aged 55 years or older in Australia. Demographic characteristics, clinical data and peripheral blood were collected to assess for the presence of CH mutations and identify potential risk factors for and clinical sequelae of CH. Investigators testing for CH were blinded to participants’ HIV status. In total, 132 CH mutations were identified in 99 (22.2%) of 446 participants. CH was more prevalent in HIV-positive participants than HIV-negative participants (27.7% vs. 16.8%, p =0.006), overall and across all age groups. HIV infection was associated with an increased odds of having CH (adjusted odds ratio 2.10, 95% confidence interval 1.30-3.38, p=0.002). The most common genes mutated were *DNMT3A* (48.5%), *TET2* (20.5%) and *ASXL1* (11.4%). CH and HIV infection were independently associated with increases in blood parameters and biomarkers associated with inflammation. These data suggest a selective advantage for the emergence of CH in the context of chronic infection and inflammation related to HIV infection.

## Introduction

As effective and well tolerated modern antiretroviral therapy (ART) regimens are becoming widely available in many settings, people with HIV who receive optimal ART are living longer^1,2^ with life expectancies approaching those of people without HIV^3-5^. However, HIV infection remains associated with an increased prevalence of comorbidities^4,6^ such as cardiovascular disease^6-8^ and non-AIDS defining malignancies^7,9^. While the reasons for this are not clear, increased comorbidities are found even in people with HIV who are virologically suppressed on ART^10,11^, suggesting additional factors such as inflammation and immune activation^11-13^, adverse effects of ART^14,15^, immunodeficiency^9,16,17^, and altered coagulation^18,19^ are playing a role. In addition, traditional risk factors such as smoking^20^ and alcohol use^21^ as well as exposure to oncogenic viruses^22^ also contribute to the increased prevalence of comorbidities in people with HIV.

Clonal haematopoiesis (CH) is characterised by the over production of blood cells derived from a single haematopoietic stem cell (HSC) harbouring somatic mutation(s) that confer a selective clonal advantage. In recent years, large studies in the general population have shown that CH increases with age and is associated with the development of haematologic malignancies, cardiovascular disease, stroke, and all-cause mortality^23-27^. While the pathophysiology of clonal dominance in CH is still largely unexplained, a number of studies have suggested an important interplay between CH and inflammation^28-30^. Despite the established causal relationship between chronic viral infections and chronic immunoinflammation, the consequence of a sustained virally-mediated underlying inflammatory state on the prevalence of CH remains unknown. Given the increased incidence of cardiovascular disease and malignancies in people with HIV and the established link between CH and cardiovascular and malignant outcomes in the general population, we sought to evaluate the prevalence of CH among persons with HIV.

The Age-Related Clonal haematopoiesis in an HIV Evaluation cohort (ARCHIVE) study was established to study genomic factors associated with ageing and the development of comorbidities among men and women with and without HIV over the age of 55 years. Our objectives in the ARCHIVE cohort were to: 1) compare the prevalence and type of somatic mutations in people with and without HIV; 2) explore socio-demographic and clinical characteristics among all participants, and characteristics of HIV infection among HIV-positive participants, in order to identify risk factors for the development of CH mutations; and 3) explore clinical outcomes including inflammatory biomarker profiles and medical comorbidities among people with and without somatic mutations, stratified by HIV status.

## Methods

### Study Design and Population

The detailed ARCHIVE study protocol is included as a supplementary appendix. In brief, the study enrolled participants over the age of 55 years from five high HIV-case load primary care practices and one tertiary hospital (a designated HIV care centre) in Sydney, Australia, and one high HIV-case load primary care practice and two tertiary hospitals (both designated HIV care centres) in Melbourne, Australia. HIV-negative participants were required to have a negative HIV test within 12 months of enrollment.

After providing written informed consent, participants provided a whole blood sample for full blood examination, inflammatory biomarker testing (Supplementary Table S1) and for CH testing. Relevant data were obtained from participants’ medical records including: age, gender, sexual orientation, most recent vital signs, results of routine standard of care blood tests (Supplementary Table S1), medical diagnoses, and use of pre-exposure prophylaxis (PrEP) among HIV-negative participants. For participants with HIV, date of HIV diagnosis, date of initiation of ART, lowest CD4 count on record, most recent CD4 count and HIV viral load, prior exposure to stavudine and zidovudine, which are known to cause myelosuppression^31^, and current ART regimen were also recorded. All participants were asked to self-complete a questionnaire on their ancestry, physical health / frailty, use of tobacco, alcohol and recreational drugs, and household income. BMI was calculated using height and weight reported from the enrolling clinical site, and was categorised using ranges published by the Centers for Disease Control and Prevention^32^. Data on sexual orientation was obtained by asking the enrolling clinical site to report whether the participant is sexually active with men, women or both. Sexual orientation categorised as men who have sex with men (MSM) was defined as male participants who were reported by the clinical site as having sex with either men only or both men and women.

### Testing for Mutations Associated with Clonal Haematopoiesis

Investigators completing the testing for CH were blinded to participants’ HIV status. DNA was obtained from whole blood from enrolled participants. Screening for clonal haematopoiesis was performed at the Peter MacCallum Cancer Centre using a bespoke targeted deep sequencing (TS) amplicon panel designed across 55 genes recurrently mutated in haematological malignancies^33^ (Supplementary Methods). Target specific amplification was then performed using the Fluidigm Access Array™ system. Identification of clonal haematopoiesis mutations through variant calling and curation was performed using an in-house pipeline^34^ as outlined in the Supplementary Methods.

### Statistical Considerations

#### Sample Size Calculation

We assumed that 10% of our HIV-negative participants would have somatic mutations associated with clonal haematopoiesis, which is consistent with published studies in the literature reporting a range of 6-10% in older adults^23-25^. Testing 220 HIV-positive and 220 HIV-negative participants gave 80% power to detect an increase to 20% prevalence of mutations among HIV-positive participants compared to the background mutation rate of 10% among HIV-negative participants.

#### Statistical Analysis

Conventional descriptive statistics were used to compare characteristics across HIV-positive and HIV-negative participants; p-values for comparisons were generated using the chi-square test. The primary outcome of interest was the presence of any somatic mutation associated with clonal haematopoiesis. We used logistic regression to calculate the unadjusted and adjusted odds ratios (OR) and 95% confidence interval (CI) for the risk of CH among HIV-positive participants, compared with HIV-negative participants. We adjusted *a priori* for age, gender and whether the participant had ever smoked, which are factors that have been reported to be associated with clonal haematopoiesis in the general population^23-25^ and with HIV infection (smoking and gender). We also adjusted for MSM status, which was found to differ slightly across HIV status in our study population.

In exploratory secondary analyses we looked for associations between HIV specific risk factors and CH using logistic regression. We also explored associations between our explanatory variables of interest (HIV infection and presence of CH) and clinical outcomes including elevations of: 1) inflammatory markers, 2) blood cell counts and characteristics and 3) select clinical comorbidities, including cardiovascular disease, malignancies and specifically, haematologic malignancies. Multivariable linear and logistic regression methods, adjusting *a priori* for age, gender, and history of ever smoking, were used to evaluate the relationship between HIV and CH and our continuous and dichotomous clinical outcomes, respectively. All continuous outcomes underwent natural log (ln) transformation. No adjustments were made to significance levels or confidence intervals for multiple comparisons. The data analysis for this paper was generated using SAS software, Version 9.4 of the SAS System for Windows (Copyright © 2019 SAS Institute Inc., Cary, NC, USA).

#### Human Subjects

The research protocol, consent form, and associated documents were approved by the relevant Human Research Ethics Committees in both the states of New South Wales and Victoria, Australia, where participants were enrolled. All study participants gave written informed consent to participate in the study.

## Results

### Study participant characteristics

A total of 461 patients were consented and 446 participants (220 HIV-positive and 226 HIV-negative) were included in the final analysis (Figure 1). Overall, HIV-positive and HIV-negative participants were similar in terms of demographic and clinical characteristics (Table 1, Supplementary Table 1). Among all participants, the median age (IQR) was 63 (59-69) years and 96.2% were male. Among the 390 (97.1%) with available data on sexual orientation from the enrolling clinical site, more HIV-positive participants were reported to be MSM than HIV-negative participants (93.1% and 87.1%, respectively). Of the 446 participants included, 397 (89.0%) completed the on-line questionnaire, including 191 (86.8%) with HIV and 206 (91.2%) without HIV. Half (50.6%) of respondents who completed the questionnaire reported ever using tobacco, though only 11.6% reported currently smoking. The majority (77.5%) of participants reported currently drinking alcohol.

**Table 1.** Demographic and clinical characteristics of study participants, overall and by HIV status

|  | HIV Negative | HIV Positive | All Participants |
| --- | --- | --- | --- |
| Characteristic | N = 226 | N = 220 | N = 446 |
|  | n (%) | n (%) | n (%) |
| **Demographics and clinical characteristics** |  |  |  |
| **Age (years)** |  |  |  |
| Median (IQR); range | 64 (59-69); 55-82 | 63 (59-68); 55-84 | 63 (59-69); 55-84 |
| 55-64 years | 118 (52.2) | 125 (56.8) | 243 (54.5) |
| 65-74 years | 91 (40.3) | 87 (39.6) | 178 (39.9) |
| ≥ 75 years | 17 (7.5) | 8 (3.6) | 25 (5.6) |
| **Gender** |  |  |  |
| Male | 213 (94.3) | 216 (98.2) | 429 (96.2) |
| Female | 13 (5.8) | 4 (1.8) | 17 (3.8) |
| **Sexual orientation** |  |  |  |
| Men sexually active with men | 189 (83.6) | 201 (91.4) | 390 (87.4) |
| Other | 28 (12.4) | 15 (6.8) | 43 (9.6) |
| Missing | 9 (4.0) | 4 (1.8) | 13 (2.9) |
| **Tobacco use** |  |  |  |
| Never smoked | 108 (47.8) | 87 (39.6) | 195 (43.7) |
| Used to smoke | 76 (33.6) | 78 (35.5) | 154 (34.5) |
| Currently smoke | 20 (8.9) | 26 (11.8) | 46 (10.3) |
| Missing | 22 (9.7) | 29 (13.2) | 51 (11.4) |
| **Alcohol use** |  |  |  |
| Never drank | 7 (3.1) | 10 (4.6) | 17 (3.8) |
| Used to drink | 40 (17.7) | 32 (14.6) | 72 (16.1) |
| Currently drink | 157 (69.5) | 149 (67.7) | 306 (68.6) |
| Missing | 22 (9.7) | 29 (13.2) | 51 (11.4) |
| **Recreational drug use in past 12 months** |  |  |  |
| No | 154 (68.1) | 135 (61.4) | 289 (64.8) |
| Yes | 47 (20.8) | 56 (25.5) | 103 (23.1) |
| Missing | 25 (11.1) | 29 (13.2) | 54 (12.1) |
| **Injection drug use in the past 12 months** |  |  |  |
| No | 195 (86.3) | 181 (82.3) | 376 (84.3) |
| Yes | 5 (2.2) | 7 (3.2) | 12 (2.7) |
| Missing | 26 (11.5) | 32 (14.6) | 58 (13.0) |
| **Frailty / physical health** |  |  |  |
| Health limits vigorous activity |  |  |  |
| Not limited at all | 64 (28.3) | 67 (30.5) | 131 (29.4) |
| Limited at least a little | 133 (58.9) | 118 (53.6) | 251 (56.3) |
| Missing | 29 (12.8) | 35 (15.9) | 65 (14.4) |
| Falls in past 12 months |  |  |  |
| None | 169 (74.8) | 148 (67.3) | 317 (71.1) |
| Any | 32 (14.2) | 39 (17.7) | 71 (15.9) |
| Missing | 25 (11.1) | 33 (15.0) | 58 (13.0) |
| **Medical conditions** |  |  |  |
| At least one condition | 222 (98.2) | 213 (96.8) | 435 (97.5) |
| Cardiovascular conditions | 148 (65.5) | 141 (64.1) | 289 (64.8) |
| Malignancies | 48 (21.2) | 60 (27.3) | 108 (24.2) |
| Liver conditions, any | 35 (15.5) | 45 (20.5) | 80 (17.9) |
| Viral Hepatitis B | 11 (4.9) | 20 (9.1) | 31 (7.0) |
| Viral Hepatitis C | 20 (8.9) | 20 (9.1) | 40 (9.0) |

**Figure 1.**
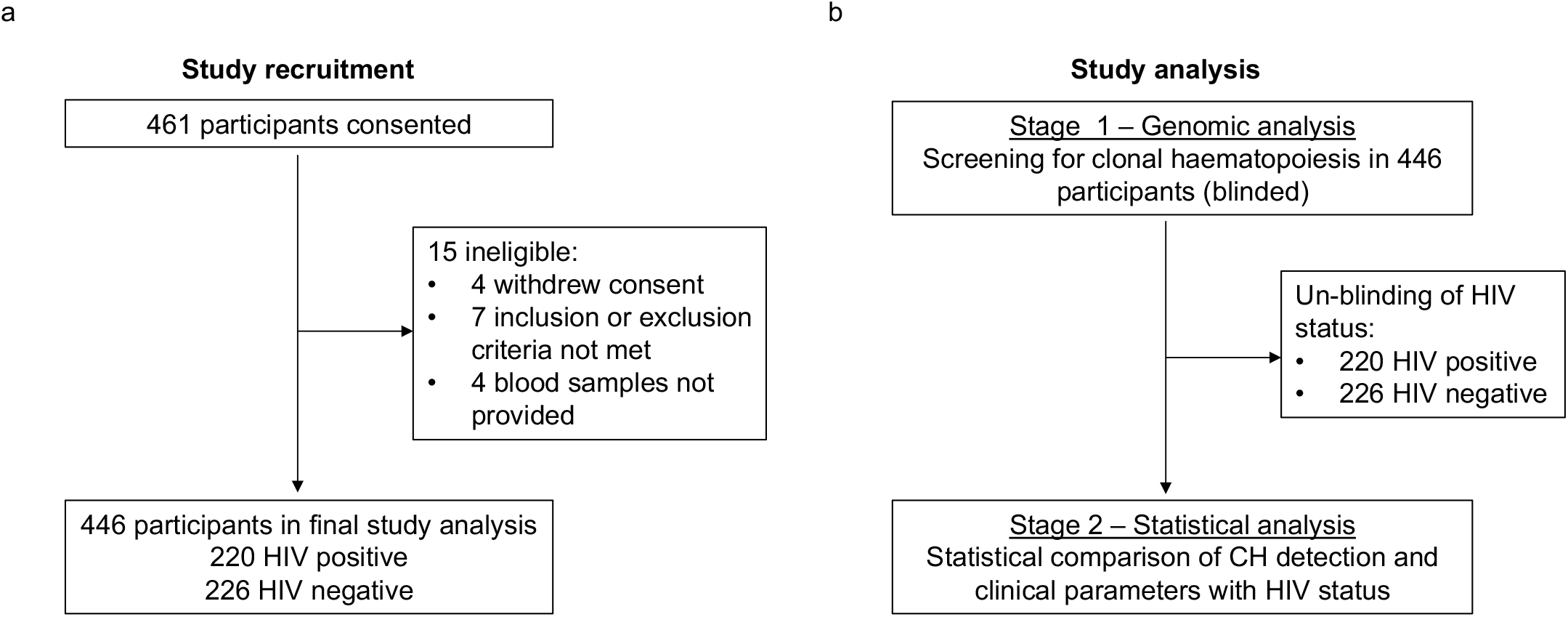
ARCHIVE study recruitment and analysis. 1a. Participant enrolment and inclusion in the final analysis 1b. Study analysis design

Vital signs at the time of enrolment, clinical laboratory results available from the medical record, and study pathology results were consistent with a generally healthy study population, although 19.5% of participants were obese, defined as a body mass index ≥30 (Table 1, Supplementary Table 1). Almost all participants (97.5%) were reported by the enrolling clinical site to have at least one medical condition, excluding HIV infection (Table 1 and Supplementary Table 1). The majority (64.8%) had cardiovascular comorbidities, with hypertension (38.3%) and hyperlipidaemia (37.7%) being the most common. Twenty-four percent of participants had a history of malignancy with the majority (61.1%) being skin cancers. HIV-positive participants had a higher prevalence of several medical comorbidities, compared to HIV-negative participants (Supplementary Table 1). HIV-positive participants were found to have lower LDL, and higher triglycerides, creatinine, haemoglobin, lymphocyte count, mean corpuscular volume, interleukin-6 (IL-6), and Cystatin C, than HIV-negative participants (Supplementary Table 1).

Among the 216 HIV-positive participants with available data on date of HIV diagnosis, the median (IQR) number of years from diagnosis to enrolment was 24 (15-31) (Supplementary Table 2). Of the 217 participants with available data on CD4 nadir, 34.6% had a CD4 nadir under 200 cells/mm^3^. Seventeen percent had a previous AIDS-defining condition; the most common were *Pneumocystis jiroveci* pneumonia (6.8%), oral candidiasis (5.9%), and Kaposi’s sarcoma (4.1%) (Supplementary Table 1). Most HIV-positive participants (69.6%) had a current CD4 count of ≥ 500 cells/mm^3^, 95.5% had a current HIV viral load less than 40 copies/mL, and 99.6% were currently taking ART. Of the 211 with available historical ART data, 19.0% initiated ART prior to 1996, when “highly active” ART became widely used in Australia, 41.7% initiated ART from 1996-2005, and 39.3% initiated ART since 2006 when modern ART regimens became available. The median (IQR) number of years since starting ART was 17 (9-21) and 38.4% had been taking ART for 20 years or more.

**Table 2.**
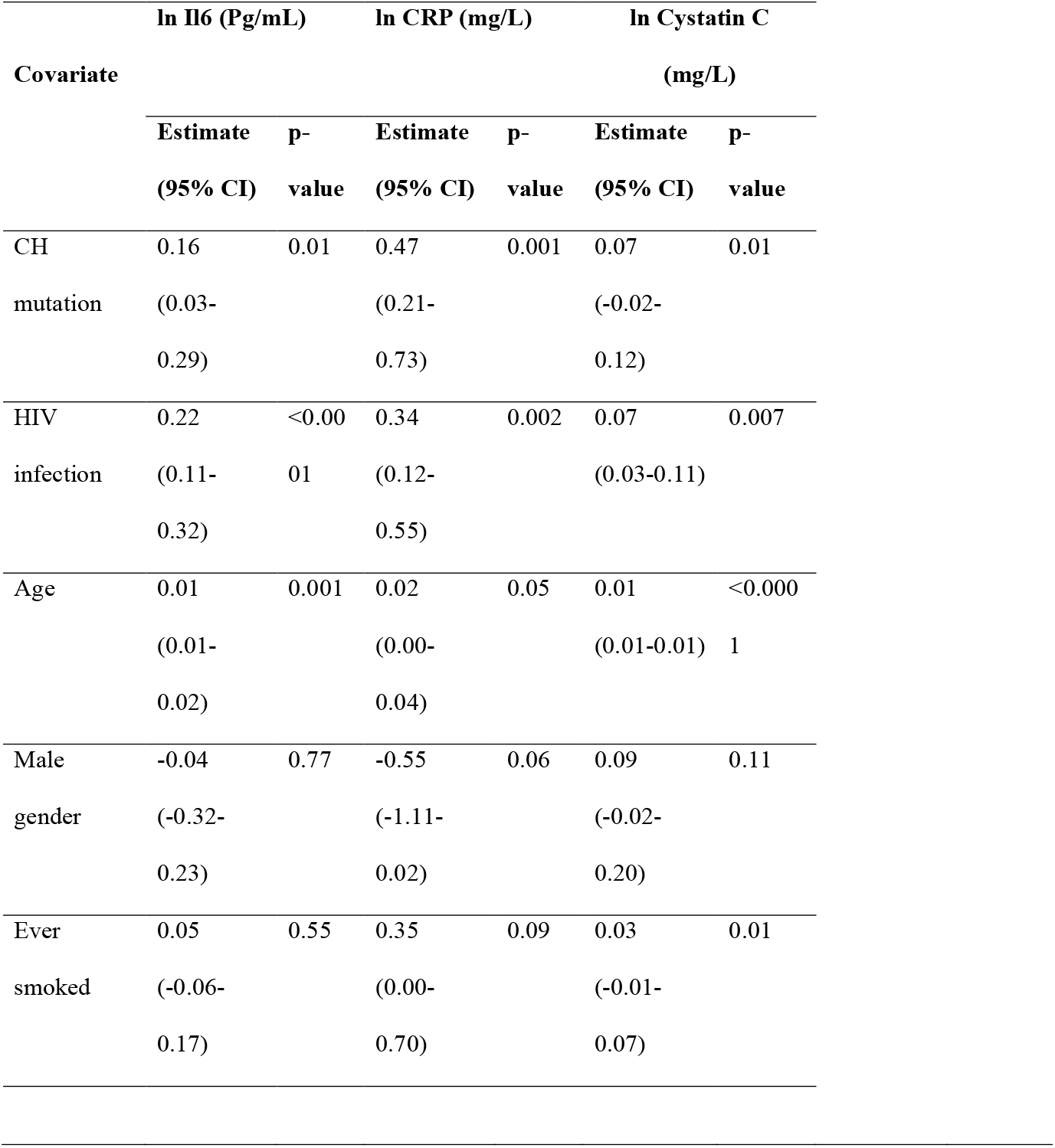
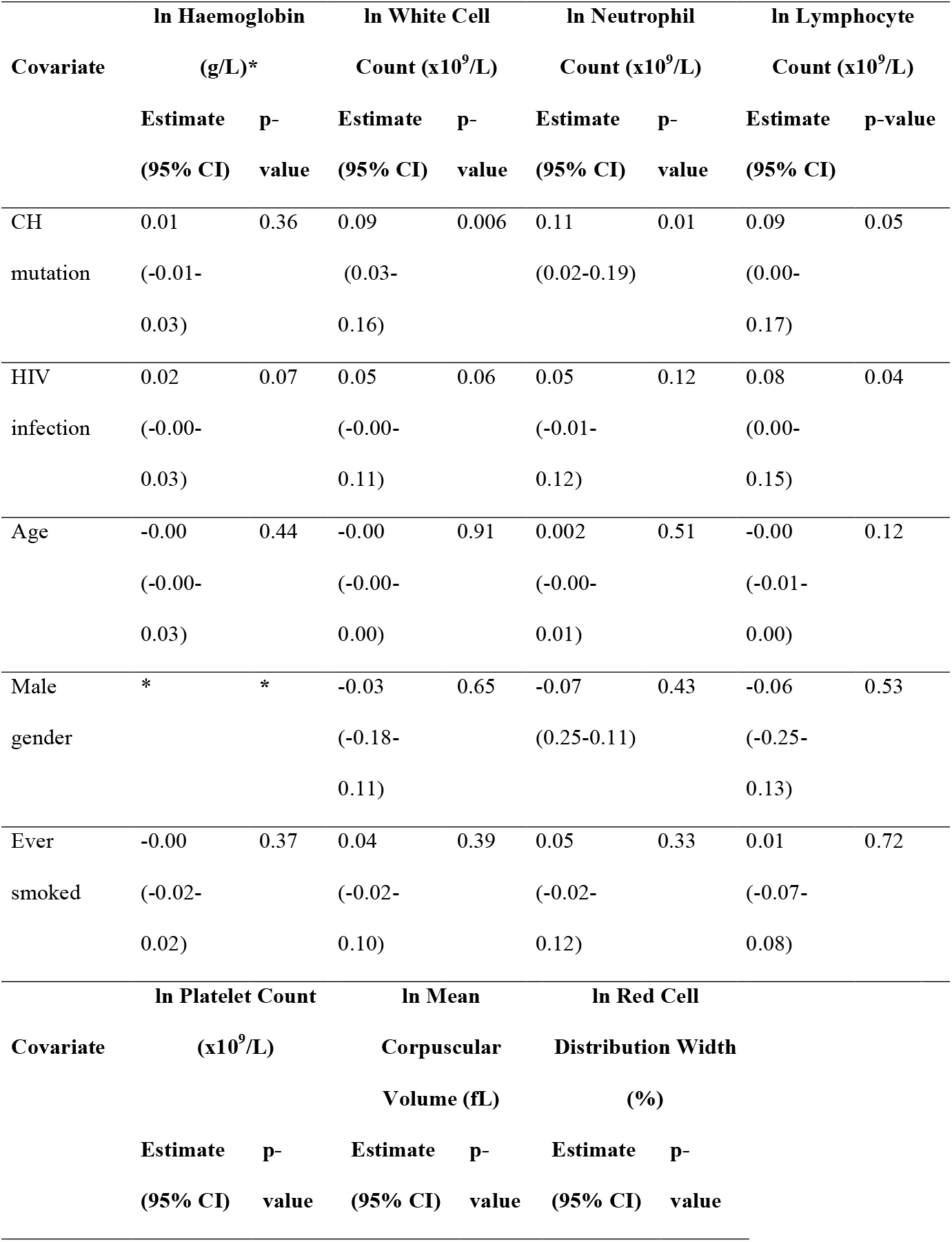
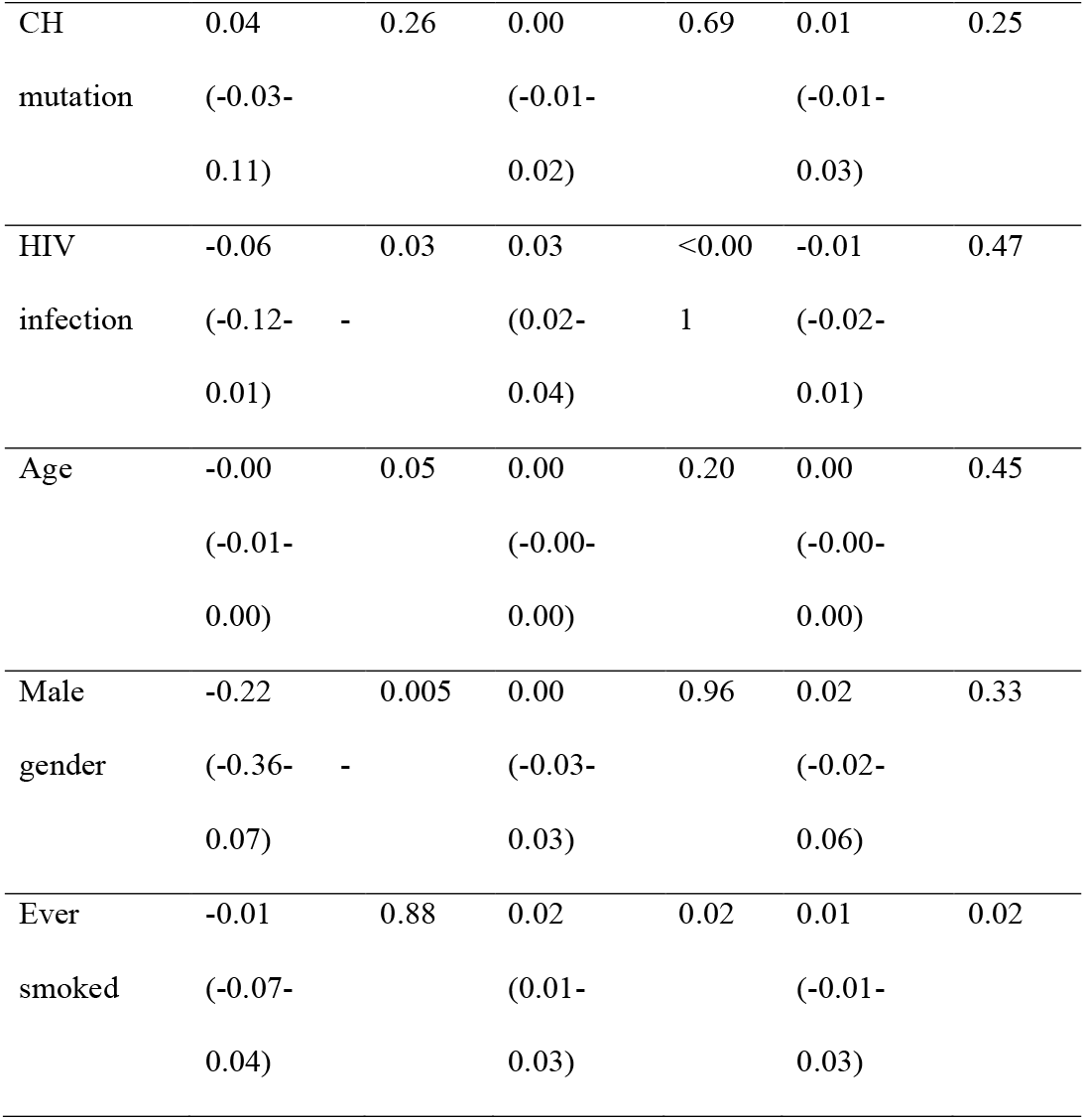
Multivariable regression analysis of associations between the presence of at least one CH mutation, HIV status, age, gender, and history of ever smoking and clinical outcomes including blood cell counts, inflammatory markers, and comorbidities **Table 2a.** Inflammatory markers and FBE (linear regression)

**Table 2b.** D-dimer and comorbidities^**1**^ (logistic regression)

| Covariate | D-dimer $\geq 0.27$ | | Cardiovascular<sup>2</sup> disease | | Malignancies (all) | | Haematologic malignancies | |
| --- | --- | --- | --- | --- | --- | --- | --- | --- |
|  | OR | p- | OR | p- | OR | p- | OR | p-value |
|  | (95% CI) | value | (95% CI) | value | (95% CI) | value | (95% CI) |  |
| CH | 1.09 | 0.73 | 0.94 | 0.86 | 1.04 | 0.90 | 0.39 | 0.39 |
| mutation | (0.68-1.73) |  | (0.50-1.78) |  | (0.61-1.76) |  | (0.04-3.33) |  |
| HIV | 0.91 | 0.63 | 1.42 | 0.21 | 1.38 | 0.16 | 8.05 | 0.05 |
| infection | (0.62-1.33) |  | (0.82-2.46) |  | (0.88-2.16) |  | (0.97-67.04) |  |
| Age | 0.95 | 0.002 | 1.10 | <0.001 | 1.03 | 0.15 | 1.02 | 0.80 |
|  | (0.92-0.98) |  | (1.05-1.15) |  | (0.99-1.07) |  | (0.90-1.16) |  |
| Male | 2.58 | 0.08 | ** | ** | 2.24 | 0.29 | ** | ** |
| gender | (0.88-7.54) |  |  |  | (0.50-10.09) |  |  |  |
| Ever | 0.82 | 0.46 | 1.59 | 0.82 | 0.85 | 0.16 | 0.68 | 0.73 |
| smoked | (0.55-1.22) |  | (0.88-2.84) |  | (0.53-1.36) |  | (0.15-3.12) |  |

OR=odds ratio; CI=confidence interval
<sup>1</sup>All comorbidities were reported by the clinic based on review of the medical record.
<sup>2</sup>Cardiovascular disease was defined as the presence of any of the following conditions: coronary artery disease, myocardial infarction, or cerebrovascular accident. Malignancies were defined as any reported cancer.
\*Women were excluded from the haemoglobin model due to varying reference ranges for men compared with women
\*\*No females in our study were reported to have cardiovascular disease or haematologic malignancies

### Identification of clonal haematopoiesis mutations

We identified a higher prevalence of CH among HIV-positive participants compared with HIV-negative participants. Overall, CH associated mutations were identified in 99 (22.2%) of all 446 study participants: 61 (27.7%) of the 220 HIV-positive participants and 38 (16.8%) of the 226 HIV-negative participants (p=0.006). The increased prevalence of CH in the HIV-positive cohort was seen across all age groups but was greatest in those over 75 years of age (Figure 2a).

**Figure 2.**
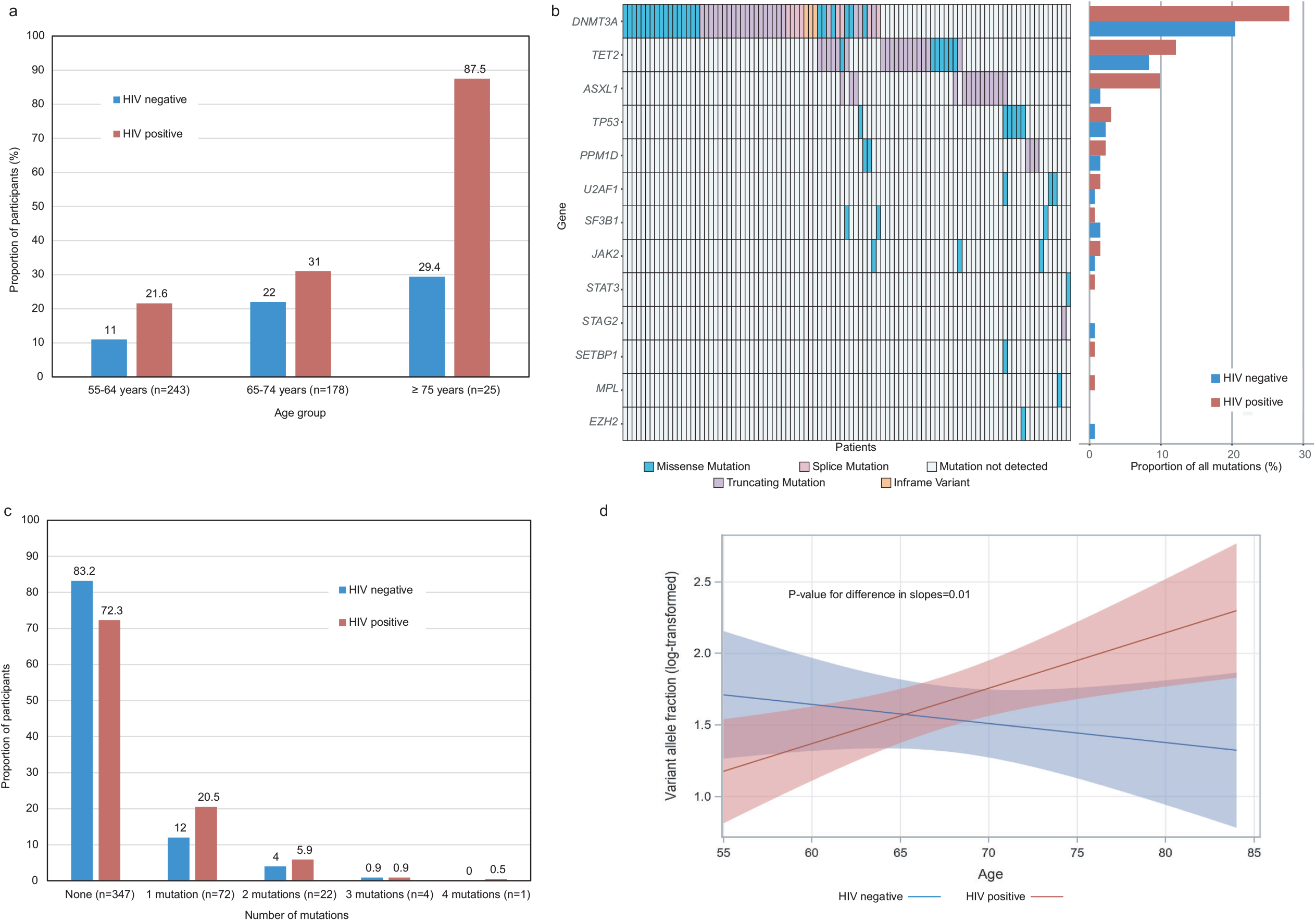
Characteristics of 132 mutations found in 99 of 446 study participants (220 HIV positive and 226 HIV negative) Figure 2a. Proportion of participants with any mutation, by age group and HIV status Figure 2b. Gene mutations in each participant (N=99 with any mutation; left panel) and proportion of mutations in each gene, by HIV status (right panel) Figure 2c. Proportion of participants with 0, 1, 2, 3 and 4 mutations, by HIV status Figure 2d. Variant allele fraction (ln-transformed) over increasing age, by HIV status

A total of 132 CH mutations were identified in the 99 participants; 81 (61.4%) in HIV-positive participants compared with 51 (38.6%) in HIV-negative participants (p =0.002). The majority occurred in three of the main genes identified in other studies of CH in the general population^23- 25^: *DNMT3A* (48.5%), *TET2* (20.5%) and *ASXL1* (11.4%). HIV-positive participants had a higher proportion of mutations in all of these genes, particularly *ASXL1*, compared with HIV-negative participants (Figure 2b). Overall, missense mutations were the most frequent mutation type (40.2%), although a higher proportion of truncating mutations was observed than previously reported^23,24^, which included frameshift (34.9%), stop-gained (15.9%), and splice site mutations (6.9%) (Figure 2b, Supplementary Table S3). Further data on mutation characteristics can be found in Supplementary Table S3 and a full list of all identified mutations associated with clonal haematopoiesis can be found in Supplementary Table S4.

More HIV-positive participants had greater than one pathogenic CH mutation detected compared to HIV-negative participants (7.3% vs 4.9%, respectively, p=0.29) (Figure 2c). In addition, the ln-transformed variant allele fractions (VAF) of the identified mutations increased with age at a greater rate among HIV-positive participants, compared with HIV-negative participants (p-value for interaction=0.01, Figure 2d); however, there was no difference in the median (IQR) VAF by HIV status (4.7 [2.6-8.6] in HIV-positive participants vs. 4.0 [2.4-8.4] in HIV-negative participants; p=0.59).

Of the 226 HIV-negative participants, 26.6% had taken PrEP to prevent HIV acquisition and of those, 50% were taking PrEP at the time of enrolment. Of the 96.7% who had available data on what PrEP agent was used, all had used tenofovir/emtricitabine. Among HIV-negative participants, there was no difference in exposure to PrEP among those with and without CH: 26.1% of HIV-negative participants without CH had ever used PrEP vs. 11 (29.0%) of those with one or more CH mutation (p=0.71).

### Association between HIV infection and the presence of clonal haematopoiesis mutations

The odds of having at least one CH mutation among HIV-positive participants compared with HIV-negative participants was 2.10 (confidence interval [CI] 1.30-3.38), p=0.002, when adjusted for age, gender, sexual orientation and history of smoking (Figure 3). In addition to fitting linear age in the model, we examined a quadratic term for age but found no statistical significance (p=0.59) We performed a sensitivity analysis of the association between HIV infection and CH by conducting four sub-analyses (mutations with VAF>2%, *DNMT3A*/*TET2*/*ASXL1* mutations, male participants, and MSM participants) and found no significant differences in our results (Supplementary Tables S5. A-D). Due to the large difference seen across HIV status in ASXL1 mutations and the association between ASXL1 and smoking^35^, we performed a sensitivity analysis excluding participants with ASXL1 mutations and did not find any significant differences in our results (data not shown).

**Figure 3.**
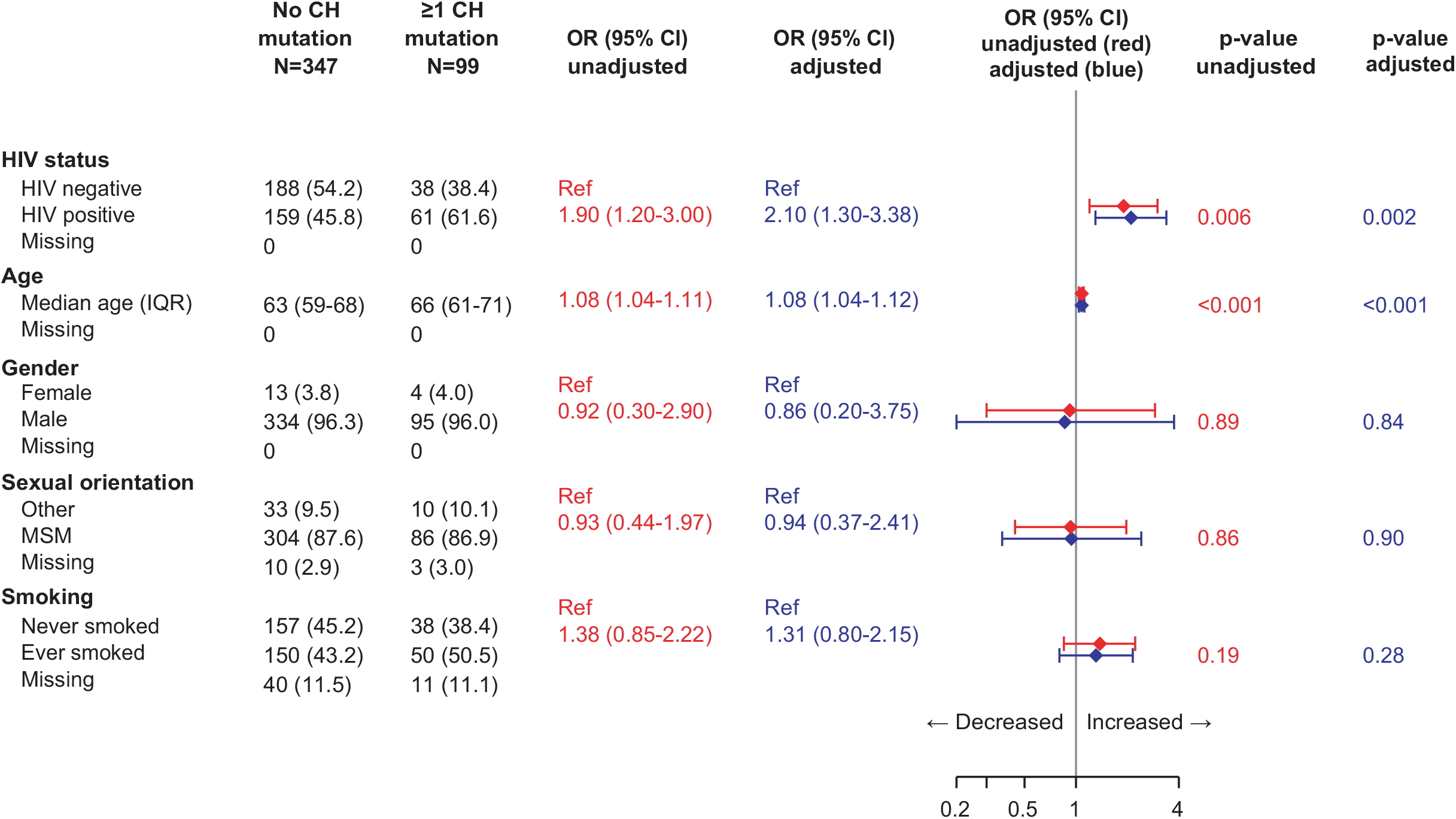
Adjusted and unadjusted estimates for the association between CH mutation and HIV status, age, gender, sexual orientation and smoking

As more HIV-positive participants were enrolled from hospital clinics than HIV-negative participants (64 [29.1%] vs. 11 [4.9%], respectively; p=<0.001), we explored whether this led to a selection bias for more unwell and possibly older, HIV-positive participants, which may have influenced our results towards finding a difference in CH by HIV status. We found no difference in the proportion of participants with and without CH that were enrolled at tertiary care sites (56 [16.1%] vs. 19 [19.2%], respectively; p=0.47) and no difference in the median age of participants enrolled at GP vs. tertiary care sites (median IQR 63 [59-69] vs. 66 [60-69]; p=0.15). In addition, we found no significant difference in the association between HIV and CH shown in Figure 3 when we adjusted for tertiary care vs. GP site in our multivariable model (data not shown).

We explored other factors that might increase the risk of CH in our study population, including ancestry, body mass index, duration and quantity of smoking exposure, alcohol use, recreational and injection drug use, and annual household income, but did not identify any statistically significant covariates (data not shown). Among HIV-positive participants, we did not identify any HIV-specific characteristics that were associated with having an increased risk of CH on univariate analysis (Supplementary Table S6). We also explored whether VAF (averaged within individuals) was correlated with years of HIV infection but did not find a significant correlation (Supplementary Figure 1).

### Association between clonal haematopoiesis and clinical outcomes

We evaluated clinical outcomes associated with both HIV infection and CH, including blood cell counts and characteristics, inflammatory markers, and select clinical comorbidities. Overall, compared to HIV-negative participants, HIV-positive participants had elevated blood parameters associated with chronic inflammation, including neutrophil count, CRP, and IL-6 (Figure 4 Panels A-C and Supplementary Figure 2 Panels A-C), even in the absence of CH. In the presence of CH, both HIV-negative and HIV-positive participants had increased levels of inflammatory biomarkers. The higher baseline levels seen in HIV-positive participants were increased even further in those with more than one CH mutation.

**Figure 4.**
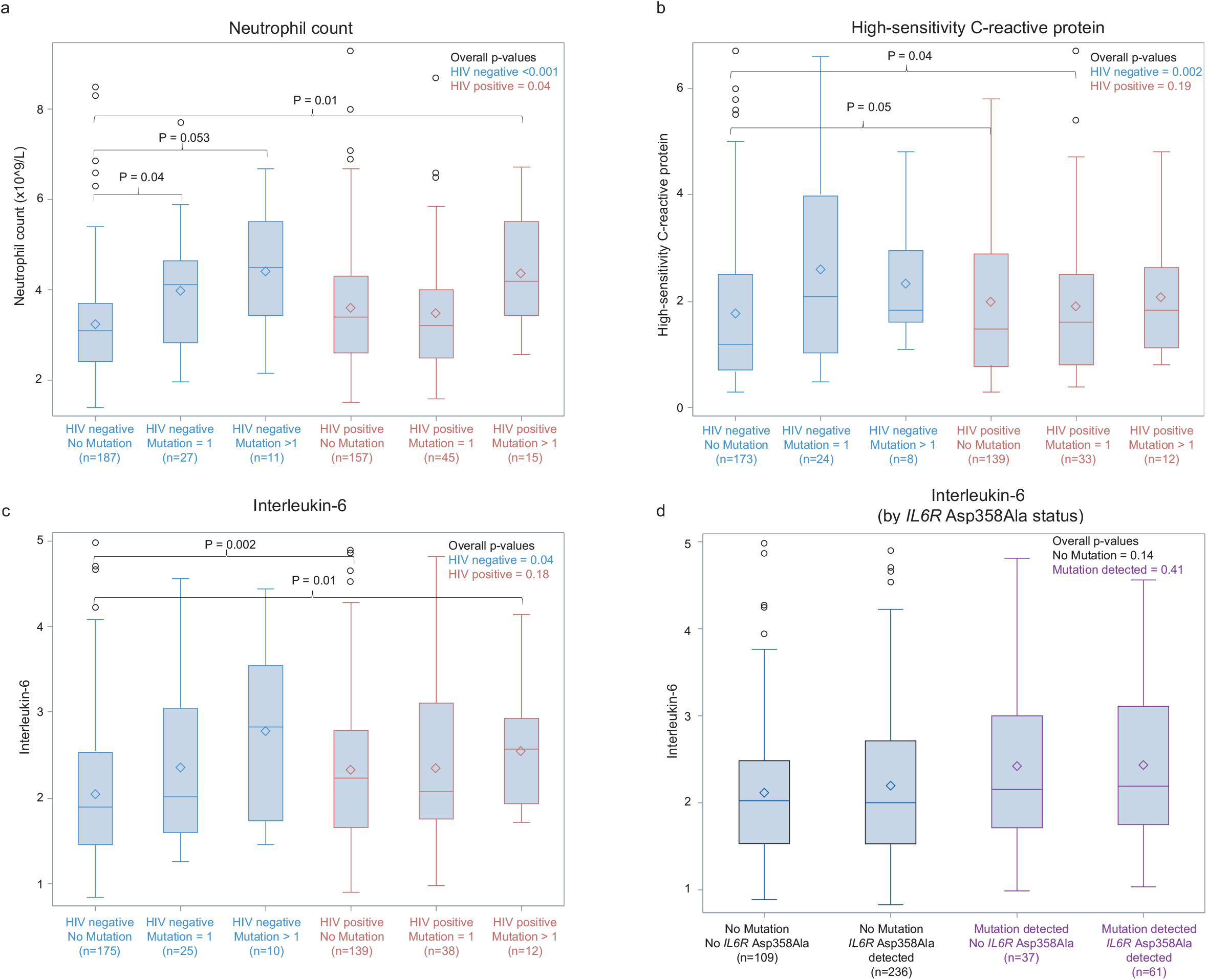
Box plots of the distribution of blood cell characteristics and inflammatory markers. Overall p-values comparing trends within subgroups (figure inset) were generated using the Kruskal-Wallis test. P-values across specific groups comparing median values (brackets) were generated using post-hoc pairwise two-sided multiple comparison analysis using the Dwass, Steel, Critchlow-Fligner Method. Panel A. Neutrophil count by HIV status and presence of 0, 1 and >1 CH mutation Panel B. High-sensitivity C-reactive protein* by HIV status and presence of 0, 1 and >1 CH mutation Panel C. Interleukin-6* by HIV status and presence of 0, 1 and >1 CH mutation Panel D. Interleukin-6* by *IL6R p. Asp358Ala* status and presence or absence of CH mutations *Outliers have been removed for box plot presentation only; p-values were calculated including outliers. Outliers were defined as values greater than 1.5 times the IQR above the third quartile.

### Interleukin-6 receptor polymorphism (IL6R p. Asp358Ala)

Recent data suggest that the common hypomorphic variant of the IL-6 receptor (*IL6R* p. Asp358Ala) potentially negates the increased risk of cardiovascular disease observed in patients with CH^36^. We found within our cohort that there was no significant difference in the prevalence of the IL6 receptor polymorphism by HIV status (62.7% among HIV-positive vs. 70.8% among HIV-negative participants, p=0.07) or across the presence of CH (61.6% among participants with CH vs. 68.3% among participants without CH, p=0.21). We also found that the increased levels of serum IL-6 noted in individuals with CH and patients with HIV infection without CH could not be explained by expression of the *IL6R* p. Asp358Ala variant (Figure 4 Panel D, Supplementary Figure 2 Panel D).

We also conducted a multivariable regression analysis of the association between the presence of CH, HIV status, and clinical outcomes including FBE characteristics, inflammatory markers and medical comorbidities, adjusting for age, gender and history of smoking, which are known confounders of our clinical outcomes (Table 2). The presence of at least one CH mutation and HIV infection were independently associated with a statistically significant increase (p<0.05) in several blood parameters associated with chronic inflammation (Table 2). However, there were little differences found in other blood count parameters including haemoglobin, platelets or red cell distribution width.

## Discussion

In this cohort of HIV-positive and HIV-negative Australian adults over the age of 55, HIV infection was associated with more than twice the odds of having CH, and this association was independent of previously reported risk factors for CH such as smoking, gender and age. Both HIV infection and the presence of CH were independently associated with increases in several parameters associated with inflammation even when adjusting for known risk factors.

We found that the increased prevalence of CH in HIV-positive individuals was most marked in older age groups. Although these individuals had higher numbers of *DNMT3A* and *TET2* mutations, the greatest difference across HIV status occurred in pathogenic mutations in *ASXL1* (Figure 2b). Notably, *ASXL1* and *DNMT3A* mutations have been identified to be more frequent in myelodysplastic syndrome (MDS) patients with HIV and are associated with a poorer prognosis^37^. Whilst MDS remains an incurable disease in the absence of an allogeneic transplantation, the impending emergence of well tolerated novel disease modifying therapies^38-40^ may argue for better surveillance and early diagnosis in older HIV-positive patients with CH.

Raised inflammatory markers, which are well-documented in HIV infection, have recently been implicated in the pathogenesis of CH^28-30,41^. Increasing experimental evidence has also highlighted the fact that repeated or sustained exposure to inflammatory stimuli are toxic to normal HSCs ultimately leading to their depletion and bone marrow failure^42-44^. Remarkably, in the context of sustained chronic inflammation, HSCs with mutations in CH genes such as TET2 have a major competitive advantage^45,46^. Unlike normal HSCs, TET2 mutant HSCs show reduced apoptosis and enhanced activity in response to inflammatory cytokines, particularly IL6^45,46^. Consistent with our findings, IL-6 is known to be increased in HIV-positive individuals even in those who are virologically-suppressed on highly effective ART regimens^19,47^. Taken together, these findings raise the prospect that the underlying chronic inflammatory state seen in HIV infection provides the ideal milieu for clonal dominance of HSC with CH mutations and may account for the increased risk of CH seen in HIV-positive individuals. It is also possible that the immunodeficiency associated with acute HIV infection and / or the chronic inflammation and immune activation associated with long-term HIV infection may lead to impaired immune surveillance and reduced clearance of clonal populations of HSC in people with HIV further contributing to the higher prevalence of CH.

Whilst these are the first prospective data from a carefully conducted clinical study designed specifically to investigate the prevalence of CH in people with and without HIV, we note with interest that a recent pre-print describing a retrospective analysis of sequencing data from disparate cohorts, including some individuals with HIV, have confirmed our findings providing independent validation of our study^48^.

Notably, our cohort only enrolled people over 55 years of age who were predominantly male and MSM in a health system that provides universal access to standard of care antiretrovirals. Further clinical studies in larger and more diverse populations including younger individuals, women with HIV and those who have not had sufficient access to ART are needed to evaluate HIV-specific risk factors for CH. These insights may inform future trial interventions to guide the management of CH in people with HIV.

## Supporting information

ARCHIVE study protocol

## Data Availability

The data that support the findings of this study are available from the corresponding authors upon reasonable request.

## Acknowledgements

The authors would like to thank all of the patients that participated in this study. We also acknowledge the following clinical site study coordinators: Trina Vincent, Holdsworth House Medical Practice; Ricardo Rosario, East Sydney Doctors; Helen Lau, Prahran Market Clinic; Florence Bascombe, St Vincent’s Hospital Sydney; Denise Smith, Albion Centre; Sally Price, Alfred Hospital; Jessica O’Brien, Monash Health; Hooi Theng Lynn Tan, Taylor Square Private Clinic; Brett Sinclair, Department of Sexual Health Medicine, Sydney Local Health District.

**Statistical analyses** were completed by N.J.D and P.Y

## References

1. Autenrieth CS, Beck EJ, Stelzle D, Mallouris C, Mahy M, Ghys P. Global and regional trends of people living with HIV aged 50 and over: Estimates and projections for 2000-2020. PLoS One 2018;13:e0207005.

2. Kirby Institute. HIV, viral hepatitis and sexually transmissible infectiosn in Australia Annual Surveillance Report 2018. Available at: https://kirby.unsw.edu.au/sites/default/files/kirby/report/KI_Annual-Surveillance-Report-2018.pdf Access date: October 31, 2020. (Accessed at

3. Gueler A, Moser A, Calmy A, et al. Life expectancy in HIV-positive persons in Switzerland: matched comparison with general population. AIDS 2017;31:427–36.

4. Marcus JL, Leyden WA, Alexeeff SE, et al. Comparison of Overall and Comorbidity-Free Life Expectancy Between Insured Adults With and Without HIV Infection, 2000-2016. JAMA Netw Open 2020;3:e207954.

5. Antiretroviral Therapy Cohort C. Life expectancy of individuals on combination antiretroviral therapy in high-income countries: a collaborative analysis of 14 cohort studies. Lancet 2008;372:293–9.

6. Schouten J, Wit FW, Stolte IG, et al. Cross-sectional comparison of the prevalence of age-associated comorbidities and their risk factors between HIV-infected and uninfected individuals: the AGEhIV cohort study. Clin Infect Dis 2014;59:1787–97.

7. Hasse B, Ledergerber B, Furrer H, et al. Morbidity and aging in HIV-infected persons: the Swiss HIV cohort study. Clin Infect Dis 2011;53:1130–9.

8. Rosenson RS, Hubbard D, Monda KL, et al. Excess Risk for Atherosclerotic Cardiovascular Outcomes Among US Adults With HIV in the Current Era. J Am Heart Assoc 2020;9:e013744.

9. Shiels MS, Engels EA. Evolving epidemiology of HIV-associated malignancies. Curr Opin HIV AIDS 2017;12:6–11.

10. Feinstein MJ, Hsue PY, Benjamin LA, et al. Characteristics, Prevention, and Management of Cardiovascular Disease in People Living With HIV: A Scientific Statement From the American Heart Association. Circulation 2019:CIR0000000000000695.

11. Deeks SG. Immune dysfunction, inflammation, and accelerated aging in patients on antiretroviral therapy. Top HIV Med 2009;17:118–23.

12. Jalbert E, Crawford TQ, D’Antoni ML, et al. IL-1Beta enriched monocytes mount massive IL-6 responses to common inflammatory triggers among chronically HIV-1 infected adults on stable anti-retroviral therapy at risk for cardiovascular disease. PLoS One 2013;8:e75500.

13. Cassol E, Malfeld S, Mahasha P, et al. Persistent microbial translocation and immune activation in HIV-1-infected South Africans receiving combination antiretroviral therapy. J Infect Dis 2010;202:723–33.

14. Friis-Moller N, Sabin CA, Weber R, et al. Combination antiretroviral therapy and the risk of myocardial infarction. N Engl J Med 2003;349:1993–2003.

15. Subbaraman R, Chaguturu SK, Mayer KH, Flanigan TP, Kumarasamy N. Adverse effects of highly active antiretroviral therapy in developing countries. Clin Infect Dis 2007;45:1093–101.

16. Chow FC, Bacchetti P, Kim AS, Price RW, Hsue PY. Effect of CD4+ cell count and viral suppression on risk of ischemic stroke in HIV infection. AIDS 2014;28:2573–7.

17. Triant VA, Regan S, Lee H, Sax PE, Meigs JB, Grinspoon SK. Association of immunologic and virologic factors with myocardial infarction rates in a US healthcare system. J Acquir Immune Defic Syndr 2010;55:615–9.

18. Kuller LH, Tracy R, Belloso W, et al. Inflammatory and coagulation biomarkers and mortality in patients with HIV infection. PLoS Med 2008;5:e203.

19. Neuhaus J, Jacobs DR, Jr., Baker JV, et al. Markers of inflammation, coagulation, and renal function are elevated in adults with HIV infection. J Infect Dis 2010;201:1788–95.

20. Tesoriero JM, Gieryic SM, Carrascal A, Lavigne HE. Smoking among HIV positive New Yorkers: prevalence, frequency, and opportunities for cessation. AIDS Behav 2010;14:824–35.

21. Freiberg MS, McGinnis KA, Kraemer K, et al. The association between alcohol consumption and prevalent cardiovascular diseases among HIV-infected and HIV-uninfected men. J Acquir Immune Defic Syndr 2010;53:247–53.

22. Pierangeli A, Antonelli G, Gentile G. Immunodeficiency-associated viral oncogenesis. Clin Microbiol Infect 2015;21:975–83.

23. Genovese G, Kahler AK, Handsaker RE, et al. Clonal hematopoiesis and blood-cancer risk inferred from blood DNA sequence. N Engl J Med 2014;371:2477–87.

24. Jaiswal S, Fontanillas P, Flannick J, et al. Age-related clonal hematopoiesis associated with adverse outcomes. N Engl J Med 2014;371:2488–98.

25. Xie M, Lu C, Wang J, et al. Age-related mutations associated with clonal hematopoietic expansion and malignancies. Nat Med 2014;20:1472–8.

26. Abelson S, Collord G, Ng SWK, et al. Prediction of acute myeloid leukaemia risk in healthy individuals. Nature 2018;559:400–4.

27. Desai P, Mencia-Trinchant N, Savenkov O, et al. Somatic mutations precede acute myeloid leukemia years before diagnosis. Nat Med 2018;24:1015–23.

28. Fuster JJ, MacLauchlan S, Zuriaga MA, et al. Clonal hematopoiesis associated with TET2 deficiency accelerates atherosclerosis development in mice. Science 2017;355:842–7.

29. Jaiswal S, Natarajan P, Silver AJ, et al. Clonal Hematopoiesis and Risk of Atherosclerotic Cardiovascular Disease. N Engl J Med 2017;377:111–21.

30. Steensma DP. Clinical consequences of clonal hematopoiesis of indeterminate potential. Blood Adv 2018;2:3404–10.

31. Micromedex. (Micromedexsolutions.com). Access date: July 25, 2020. (Accessed at

32. Defining Adult Overweight and Obesity. Centers for Disease Control and Prevention. (https://www.cdc.gov/obesity/adult/defining.html) Access date: July 31, 2020. (Accessed at

33. Yeh P, Dickinson M, Ftouni S, et al. Molecular disease monitoring using circulating tumor DNA in myelodysplastic syndromes. Blood 2017;129:1685–90.

34. Wong SQ, Waldeck K, Vergara IA, et al. UV-Associated Mutations Underlie the Etiology of MCV-Negative Merkel Cell Carcinomas. Cancer Res 2015;75:5228–34.

35. Bolton KL, Ptashkin RN, Gao T, et al. Cancer therapy shapes the fitness landscape of clonal hematopoiesis. Nat Genet 2020;52:1219–26.

36. Bick AG, Pirruccello JP, Griffin GK, et al. Genetic Interleukin 6 Signaling Deficiency Attenuates Cardiovascular Risk in Clonal Hematopoiesis. Circulation 2020;141:124–31.

37. Kaner JD, Thibaud S, Jasra S, et al. HIV portends a poor prognosis in myelodysplastic syndromes. Leukemia & lymphoma 2019;60:3529–35.

38. Sallman DA, Asch AS, Al Malki MM, et al. The First-in-Class Anti-CD47 Antibody Magrolimab (5F9) in Combination with Azacitidine Is Effective in MDS and AML Patients: Ongoing Phase 1b Results. Blood 2019;134:569-.

39. Borate U, Esteve J, Porkka K, et al. Phase Ib Study of the Anti-TIM-3 Antibody MBG453 in Combination with Decitabine in Patients with High-Risk Myelodysplastic Syndrome (MDS) and Acute Myeloid Leukemia (AML). Blood 2019;134:570-.

40. Fenaux P, Platzbecker U, Mufti GJ, et al. Luspatercept in Patients with Lower-Risk Myelodysplastic Syndromes. New England Journal of Medicine 2020;382:140–51.

41. Busque L, Sun M, Buscarlet M, et al. High-sensitivity C-reactive protein is associated with clonal hematopoiesis of indeterminate potential. Blood Adv 2020;4:2430–8.

42. Pietras EM, Mirantes-Barbeito C, Fong S, et al. Chronic interleukin-1 exposure drives haematopoietic stem cells towards precocious myeloid differentiation at the expense of self-renewal. Nat Cell Biol 2016;18:607–18.

43. Pietras EM, Lakshminarasimhan R, Techner JM, et al. Re-entry into quiescence protects hematopoietic stem cells from the killing effect of chronic exposure to type I interferons. J Exp Med 2014;211:245–62.

44. Bogeska R, Kaschutnig P, Fawaz M, et al. Hematopoietic stem cells fail to regenerate following inflammatory challenge. bioRxiv 2020:2020.08.01.230433.

45. Cai Z, Kotzin JJ, Ramdas B, et al. Inhibition of Inflammatory Signaling in Tet2 Mutant Preleukemic Cells Mitigates Stress-Induced Abnormalities and Clonal Hematopoiesis. Cell Stem Cell 2018;23:833–49 e5.

46. Meisel M, Hinterleitner R, Pacis A, et al. Microbial signals drive pre-leukaemic myeloproliferation in a Tet2-deficient host. Nature 2018;557:580–4.

47. Grund B, Baker JV, Deeks SG, et al. Relevance of Interleukin-6 and D-Dimer for Serious Non-AIDS Morbidity and Death among HIV-Positive Adults on Suppressive Antiretroviral Therapy. PLoS One 2016;11:e0155100.

48. Alexander AG, Popadin K, Thorball CW, Uddin M, Zanni M, Yu B, Cavassini M, Rauch A, Tarr P, Schmid P, Bernasconi E, Günthard HF, Libby P, Boerwinkle E, McLaren PJ, Ballantyne CM, Grinspoon S, Natarajan P, Fellay J, the Swiss HIV Cohort Study. Increased CHIP Prevalence Amongst People Living with HIV. Available at: https://www.medrxiv.org/content/10.1101/2020.11.06.20225607v1. Access date: November 11, 2020.

