## Supplemental Material for "Age-related clonal haematopoiesis is more prevalent in older adults with HIV: the ARCHIVE study"

### Supplementary Material

#### Supplementary Methods

**Genes used in next generation panel for targeted sequencing (TS).**

| GENE | | | | | | |
|---|---|---|---|---|---|---|
| ASXL1* | DIS3 | GNAS | MLL2 | PHF6 | SF3B1 | TP53 |
| BCOR | DNMT3A | IDH1 | MPL | PRPF40B | SMC1a | U2AF1 |
| BRAF | DOCK4 | IDH2 | MYC | PTEN | SMC3 | UMODL1 |
| CALR | ETV6 | JAK2 | NF1 | PTPN11 | SRSF2 | WAC |
| CBL | EZH2 | KDM6A | NPM1 | RAD21 | STAG2 | WT1 |
| CDKN2A | FLT3 | KIT | NRAS | RUNX1 | STAT3 | ZRSR2 |
| CSF1R | GATA1 | KRAS | PDGFRA | SETBP1 | SUZ12 | PPM1D |
| CUX1 | GATA2 | MLL | PDS5B | SF3A1 | TET2 | |

* Excluding *ASXL1* c.1934dupG;p.Gly646TrpfsX12.

**Whole blood sample processing**

Whole blood from enrolled participants were collected in ethylenediaminetetraacetic acid (EDTA) tubes. DNA from whole blood was extracted using the QIAsymphony DNA Midi Kit (Qiagen, #931255) according to manufacturer's protocols.

**Targeted deep sequencing**

Target specific amplification was performed using the Fluidigm Access Array™ system. Following amplification, products were harvested, tagged with sample-specific barcodes, pooled together, and purified using AMPure XP beads. All samples were analysed in duplicate to control for PCR artefacts. The purified libraries were then sequenced using 150 bp read length on the

Illumina MiSeq or NextSeq platform. Sequenced reads were mapped to the human reference genome (version hg19) using BWA-MEM (version 0.7.12) with default parameters. All samples were tested in at least 2 technical replicates. Variant calling was performed using an in-house pipeline [1]. Mutations with at least 300x coverage with a minimum of 30 reads supporting the variant and a mutant allele fraction greater than 1% were retained for further analysis. The median targeted sequencing coverage over variant reads was 3210-fold. Variants that were recurrently observed in more than 1% of the samples (representing likely sequencing and/or PCR artefacts) and those with a high global allele frequency (>1%) in population databases (gnomAD) were flagged and removed from this curated list. In addition, the *ASXL1* c.1934dupG; p.Gly646TrpfsX12 variant was excluded from analysis to eliminate the possibility false positive results given the high susceptibility to PCR artefact within this region. Variants in the curated list were then annotated based on their prognostic or functional relevance[2]. Our analysis did not include paired germline sample to definitively exclude germline variants. To account for this, variants greater than 30% allele frequency were not included in the final curated list unless they had been described in COSMIC.

**Droplet Digital PCR**

Digital PCR was used to assess for the presence of the IL6R p.Asp358Ala polymorphism. In addition, it was used to validate hotspot mutations in *DNMT3A* Arg882His, Arg882Cys and *JAK2* Val617Phe detected by targeted sequencing. This was performed using the Bio-Rad QX200 Droplet digital PCR system following manufacturer's protocols. Allele-specific PCR assays to specifically detect and quantify the fractional abundance of point mutations and corresponding wild-type alleles were commercially obtained (PrimePCR™ PCR Primers and Assays, BioRad

Laboratories). Each sample was analysed by at least two technical replicates. Data analysis was carried out using the QuantaSoft Software (Bio-Rad).

#### Supplementary Table S1. Demographic characteristics, medical conditions, and clinical pathology results of study participants

| Demographics | HIV Negative N = 226 n (%) | HIV Positive N = 220 n (%) | All Participants N = 446 n (%) |
|---|---|---|---|
| **Aboriginal or Torres Strait Islander** | | | |
| Yes | 1 (0.4) | 2 (0.9) | 3 (0.7) |
| No | 204 (90.3) | 189 (85.9) | 393 (88.1) |
| Missing | 21 (9.3) | 29 (13.2) | 50 (11.2) |
| **Ancestry** | | | |
| Australia, New Zealand, Europe, United States of America | 185 (81.9) | 168 (76.4) | 353 (79.2) |
| Other | 18 (8.0) | 22 (10.0) | 40 (9.0) |
| Missing | 23 (10.2) | 30 (13.6) | 53 (11.9) |
| **Sexual orientation** | | | |
| Men sexually active with men or both men and women (MSM) | 189 (83.6) | 201 (91.4) | 390 (87.4) |
| Heterosexual men | 16 (7.1) | 11 (5.0 | 27 (6.1) |
| Heterosexual women | 11 (4.9) | 4 (1.8) | 15 (3.4) |
| Bisexual women | 1 (0.4) | 0 | 1 (0.2) |
| Missing | 9 (4.0) | 4 (1.8) | 13 (2.9) |

| | | | |
|---|---|---|---|
| **Tobacco use** | | | |
| Number of years smoking cigarettes regularly | | | |
| Never smoked | 108 (47.8) | 87 (39.6) | 195 (43.7) |
| <30 years | 50 (22.1) | 45 (20.5) | 95 (21.3) |
| ≥30 years | 44 (19.5) | 56 (25.5) | 100 (22.4) |
| Smoked but missing data on duration | 2 (0.9) | 3 (1.4) | 5 (1.2) |
| Missing tobacco use history | 22 (9.7) | 29 (13.2) | 51 (11.4) |
| Number of cigarettes per day | | | |
| Never smoked | 108 (47.8) | 87 (39.6) | 195 (43.7) |
| <10/day | 23 (10.2) | 19 (8.6) | 42 (9.4) |
| ≥10/day | 71 (31.4) | 82 (37.3) | 153 (34.3) |
| Smoked but missing data on quantity | 2 (0.9) | 3 (1.4) | 5 (1.1) |
| Missing tobacco use history | 22 (9.7) | 29 (13.2) | 51 (11.4) |
| **Alcohol use** | | | |
| Number of years drank regularly | | | |
| Never drank | 7 (3.1) | 10 (4.6) | 17 (3.8) |
| <20 years | 32 (14.2) | 29 (13.2) | 61 (13.7) |
| ≥20 years | 161 (7.2) | 146 (66.4) | 307 (68.8) |
| Drank but missing data on duration | 4 (1.8) | 6 (2.7) | 10 (2.2) |
| Missing alcohol use history | 22 (9.7) | 29 (13.2) | 51 (11.4) |
| Average number of drinks per week | | | |

| | | | |
|---|---|---|---|
| Never drank | 7 (3.1) | 10 (4.6) | 17 (3.8) |
| ≤14 | 148 (65.5) | 162 (73.6) | 310 (69.5) |
| >14 | 46 (20.4) | 17 (7.7) | 63 (14.1) |
| Drank but missing data on quantity | 3 (1.3) | 2 (0.9) | 5 (1.1) |
| Missing alcohol use history | 22 (9.7) | 29 (13.2) | 51 (11.4) |
| **Socioeconomic factors** | | | |
| Yearly household income | | | |
| <50k | 64 (28.3) | 100 (45.5) | 164 (36.8) |
| ≥50k | 118 (52.2) | 66 (30.0) | 184 (41.3) |
| Missing | 44 (19.5) | 54 (24.6) | 98 (22.0) |
| **Medical conditions** | | | |
| Cardiovascular conditions, any | 148 (65.5) | 141 (64.1) | 289 (64.8) |
| Hypertension | 85 (37.6) | 86 (39.1) | 171 (38.3) |
| Hyperlipidaemia | 92 (40.7) | 76 (34.6) | 168 (37.7) |
| Coronary artery disease | 19 (8.4) | 25 (11.4) | 44 (9.9) |
| Myocardial infarction | 13 (5.8) | 13 (5.9) | 26 (5.8) |
| Cerebrovascular accident | 4 (1.8) | 9 (4.1) | 13 (2.9) |
| Congestive heart failure | 3 (1.3) | 1 (0.5) | 4 (0.9) |
| Other heart disease | 22 (9.7) | 26 (11.8) | 48 (10.8) |
| Coronary artery disease or myocardial infarction | 27 (12.0) | 31 (14.1) | 58 (13.0) |
| Coronary artery disease or myocardial infarction or cerebrovascular accident | 30 (13.3) | 38 (17.3) | 68 (15.3) |
| Malignancies, any | 48 (21.2) | 60 (27.3) | 108 (24.2) |

| | | | |
|---|---|---|---|
| Solid tumors other than skin cancers[1] | 20 (8.9) | 29 (13.2) | 49 (11.0) |
| Prostate | 10 (4.4) | 7 (3.2) | 17 (3.8) |
| Kaposi's sarcoma | 1 (0.4) | 9 (4.1) | 10 (2.2) |
| Anal | 2 (0.9) | 7 (3.2) | 9 (2.0) |
| Bowel | 2 (0.9) | 3 (1.4) | 5 (1.1) |
| Lung | 2 (0.9) | 0 | 2 (0.5) |
| Breast | 1 (0.4) | 0 | 1 (0.2) |
| Cervical or ovarian | 0 | 0 | 0 |
| Other[2] | 4 (1.8) | 8 (3.6) | 12 (2.7) |
| Skin cancers | 33 (14.6) | 33 (15.0) | 66 (14.8) |
| Haematologic[3] | 1 (0.4) | 7 (3.2) | 8 (1.8) |
| Liver conditions, any | 35 (15.5) | 45 (20.5) | 80 (17.9) |
| Liver conditions other than viral hepatitis | 9 (4.0) | 11 (5.0) | 20 (4.5) |
| Diabetes | 29 (12.8) | 31 (14.1) | 60 (13.5) |
| Respiratory conditions, any | 40 (17.7) | 52 (23.6) | 92 (20.6) |
| Asthma | 27 (12.0) | 29 (13.2) | 56 (12.6) |
| Community-acquired pneumonia | 4 (1.8) | 9 (4.1) | 13 (2.9) |
| Other pneumonia | 0 | 7 (3.2) | 7 (1.6) |
| Other lung condition | 15 (6.6) | 22 (10.0) | 37 (8.3) |
| Kidney disease | 7 (3.1) | 17 (7.7) | 24 (5.4) |
| Blood disorders | 12 (5.3) | 22 (10.0) | 34 (7.6) |
| Musculoskeletal, any | 66 (29.2) | 72 (32.7) | 138 (30.9) |
| Osteoporosis or other bone disease | 24 (10.6) | 33 (15.0) | 57 (12.8) |
| Hip fracture | 0 | 1 (0.5) | 1 (0.2) |

| | | | |
|---|---|---|---|
| Fracture of any bone (other than hip) | 19 (8.4) | 32 (14.6) | 51 (11.4) |
| Arthritis | 34 (15.0) | 21 (9.6) | 55 (12.3) |
| Neuropsychiatric conditions, any | 67 (29.7) | 91 (41.4) | 158 (35.4) |
| Depression | 54 (23.9) | 71 (32.3) | 125 (28.0) |
| Anxiety | 27 (12.0) | 29 (13.2) | 56 (12.6) |
| Schizophrenia or Bipolar Disorder | 5 (2.2) | 7 (3.2) | 12 (2.7) |
| Dementia including Alzheimer's Disease | 0 | 11 (5.0) | 11 (2.5) |
| Parkinson's Disease | 3 (1.3) | 2 (0.9) | 5 (1.1) |
| Neuropathy | 8 (3.5) | 40 (18.2) | 48 (10.8) |
| Shingles | 6 (2.7) | 27 (12.3) | 33 (7.4) |
| Opportunistic Infections, any | 0 | 37 (16.8) | 37 (8.3) |
| *Pneumocystis jiroveci* pneumonia | 0 | 15 (6.8) | 15 (3.4) |
| Oral candidiasis | 0 | 13 (5.9) | 13 (2.9) |
| Kaposi's sarcoma | 0 | 9 (4.1) | 9 (2.0) |
| Cryptococcal disease | 0 | 5 (2.3) | 5 (1.1) |
| Cytomegalovirus disease | 0 | 5 (2.3) | 5 (1.1) |
| Mycobacterial infection | 0 | 3 (1.4) | 3 (0.7) |
| Toxoplasmosis | 0 | 2 (0.9) | 2 (0.5) |
| Progressive multifocal leukoencephalopathy | 0 | 1 (0.5) | 2 (0.2) |

| **Vital signs**[4] | **HIV Negative N = 226 Median (IQR)** | **HIV Positive N = 220 Median (IQR)** | **All Participants N = 446 Median (IQR)** |
|---|---|---|---|
| BMI | 26.4 (23.7-29.1) | 25.7 (23.5-28.4) | 26.0 (23.6-29.0) |
| Underweight (BMI<18.5), n (%) | 1 (0.4) | 2 (0.9) | 3 (0.7) |

| | | | |
|---|---|---|---|
| Normal (BMI 18.5-<25), n (%) | 82 (36.3) | 86 (39.1) | 168 (37.7) |
| Overweight (BMI 25-<30), n (%) | 93 (41.2) | 94 (42.7) | 187 (41.9) |
| Obese (BMI ≥30), n (%) | 49 (21.7) | 38 (17.3) | 87 (19.5) |
| Missing, n (%) | 1 (0.4) | 0 | 1 (0.2) |
| Waist circumference (cm) | 99 (92-105) | 97 (91-104) | 98 (91-105) |
| Missing, n (%) | 38 (16.8) | 22 (10.0) | 60 (13.5) |
| Systolic blood pressure (mmHg) | 130 (121-140) | 130 (120-140) | 130 (121-140) |
| Missing, n (%) | 1 (0.4) | 0 | 1 (0.2) |
| Diastolic blood pressure (mmHG) | 78 (72-86) | 80 (72-85) | 79 (72-85) |
| Missing, n (%) | 1 (0.4) | 0 | 1 (0.2) |

| **Clinical pathology results** | **HIV Negative N = 226 Median (IQR)** | **HIV Positive N = 220 Median (IQR)** | **All Participants N = 446 Median (IQR)** |
|---|---|---|---|
| Total cholesterol | 4.8 (4.2-5.6) | 4.7 (4.0-5.5) | 4.7 (4.1-5.6) |
| Recency of test (days) | 83 (0-266) | 64 (0-183) | 70 (0-244) |
| Missing, n (%) | 15 (6.6) | 1 (0.5) | 16 (7.3) |
| LDL | 2.8 (2.1-3.5) | 2.5 (1.9-3.2) | 2.6 (2.0-3.4) |
| Recency of test (days) | 73 (0-264) | 76 (4-215) | 75 (0-251) |
| Missing, n (%) | 22 (9.7) | 7 (3.2) | 29 (13.2) |
| HDL | 1.3 (1.1-1.7) | 1.3 (1.0-1.5) | 1.3 (1.1-1.6) |
| Recency of test (days) | 82 (0-260) | 65 (4-191) | 70 (0-245) |
| Missing, n (%) | 22 (9.7) | 3 (1.4) | 25 (11.4) |
| Triglycerides | 1.3 (0.9-1.9) | 1.5 (1.1-2.4) | 1.4 (1.0-2.2) |
| Recency of test (days) | 72 (0-263) | 64 (0-183) | 65 (0-242) |

| | | | |
|---|---|---|---|
| Missing, n (%) | 17 (7.5) | 1 (0.5) | 18 (8.2) |
| Glucose | 5.4 (4.9-5.8) | 5.2 (4.8-6.0) | 5.3 (4.9-5.9) |
| Recency of test (days) | 89 (0-317) | 57 (0-202) | 71 (0-253) |
| Missing, n (%) | 14 (6.2) | 10 (4.5) | 24 (10.9) |
| Creatinine | 83 (74-93) | 92 (80-105) | 87 (76-99) |
| Recency of test (days) | 40 (0-219) | 15 (0-85) | 25 (0-126) |
| Missing, n (%) | 5 (2.2) | 0 | 5 (2.3) |
| ALT | 26 (19-34) | 28 (20-39) | 27 (20-36) |
| Recency of test (days) | 47 (0-244) | 18 (0-91) | 29 (0-133) |
| Missing, n (%) | 9 (3.9) | 0 | 9 (4.1) |
| AST | 25 (21-30) | 26 (22-34) | 25 (21-31) |
| Recency of test (days) | 51 (0-245) | 36 (0-122) | 42 (0-153) |
| Missing, n (%) | 10 (4.4) | 21 (9.5) | 31 (14.1) |

| **Study pathology results[5]** | **HIV Negative N = 226 Median (IQR)** | **HIV Positive N = 220 Median (IQR)** | **All Participants N = 446 Median (IQR)** |
|---|---|---|---|
| **Full blood examination** | | | |
| Haemoglobin | 146 (136-153) | 149 (141-156) | 147 (139-155) |
| Missing, n (%) | 1 (0.4) | 3 (1.4) | 4 (0.9) |
| White blood cell count | 5.7 (4.8-6.6) | 6.1 (4.8-7.7) | 5.9 (4.8-7.0) |
| Missing, n (%) | 1 (0.4) | 3 (1.4) | 4 (0.9) |
| Neutrophil count | 3.2 (2.5-4.0) | 3.4 (2.6-4.3) | 3.3 (2.5-4.2) |
| Missing, n (%) | 1 (0.4) | 3 (1.4) | 4 (0.9) |
| Lymphocyte count | 1.7 (1.4-2.1) | 1.8 (1.5-2.3) | 1.8 (2.2-1.4) |

| | | | |
|---|---|---|---|
| Missing, n (%) | 1 (0.4) | 3 (1.4) | 4 (0.9) |
| Platelets | 209 (180-240) | 200 (165-239) | 204 (175-140) |
| Missing, n (%) | 1 (0.4) | 6 (2.7) | 7 (1.6) |
| Mean Corpuscular Volume | 92 (89-95) | 95.7 (92.0-99.0) | 94 (90-97) |
| Missing, n (%) | 1 (0.4) | 3 (1.4) | 4 (0.9) |
| Red cell distribution width | 13.4 (12.9-13.9) | 13.3 (12.9-13.9) | 13.4 (12.9-13.9) |
| Missing, n (%) | 1 (0.4) | 3 (1.4) | 4 (0.9) |
| **Inflammatory markers** | | | |
| Interleukin 6 | 1.99 (1.50-2.80) | 2.37 (1.82-3.36) | 2.15 (1.61-3.03) |
| Missing, n (%) | 1 (0.4) | 2 (0.9) | 3 (0.7) |
| hs CRP | 1.5 (0.7-3.0) | 2.0 (0.9-4.1) | 1.7 (0.8-3.7) |
| Missing, n (%) | 8 (3.5) | 2 (0.9) | 10 (2.2) |
| Cystatin C | 0.93 (0.82-1.07) | 0.99 (0.88-1.15) | 0.95 (0.86-1.11) |
| Missing, n (%) | 2 (0.9) | 1 (0.5) | 3 (0.7) |
| D-dimer | | | |
| <0.27 mg/L, n (%) | 104 (46.0) | 100 (45.5) | 204 (45.7) |
| ≥0.27 mg/L, n (%) | 120 (53.1) | 116 (52.7) | 236 (52.9) |
| Missing, n (%) | 2 (0.9) | 4 (1.8) | 6 (1.4) |

[1]Other solid tumor sites include: HIV negative (n=1 for all): kidney, thyroid, sarcoma, tongue; HIV positive (one participant had two cancers): bladder (n=2), facial nerve (n=1), head and neck (n=1), pancreatic (n=1), liposarcoma (n=1), tongue (n=1), testicular (n=1), thyroid (n=1)

[2]Includes Kaposi's sarcoma

[3]Haematologic malignancies and years of first diagnosis: HIV negative (n=1): Non-Hodgkins Lymphoma (2003); HIV positive (n=1 for all): Hodgkins Lymphoma (1990), Hodgkins Lymphoma (2004), Non-Hodgkin's Lymphoma (2004), Hodgkin's Lymphoma (2006), Multiple Myeloma (2008), gastric

lymphoma (2011), Chronic Myeloid Leukemia (2017). One of the eight participants with haematologic malignancies also had a mutation associated with clonal haematopoiesis.

[4]The median time from vital sign measurement to enrolment was 0 days (IQR 0-0 days) overall and when stratified by HIV status

[5]Tested at study enrolment visit

#### Supplementary Table S2. Clinical characteristics of study participants with HIV

| Characteristic | N=220<br>n (%) |
|---|---|
| Years since HIV diagnosis*, median (IQR) | 24 (15-31) |
| <10 | 21 (9.6) |
| 10-19 | 64 (29.1) |
| 20-29 | 65 (29.6) |
| ≥30 | 66 (30.0) |
| Missing | 4 (1.8) |
| CD4 nadir, median (IQR) | 248 (151-383) |
| <50 | 18 (8.2) |
| 50-199 | 57 (25.9) |
| 200-349 | 76 (34.6) |
| 350-499 | 38 (17.3) |
| ≥500 | 28 (12.7) |
| Missing | 3 (1.4) |
| Current CD4, median (IQR) | 649 (464-859) |
| <200 | 4 (1.8) |
| 200-349 | 22 (10.0) |
| 350-499 | 39 (17.7) |
| ≥500 | 153 (69.6) |
| Missing | 2 (0.9) |
| Current HIV VL | |

| | |
|---|---|
| <40 | 210 (95.5) |
| 40-199 | 8 (3.6) |
| ≥200 | 2 (0.9) |
| Missing | 0 |
| Currently taking ART | |
| Yes | 219 (99.6) |
| Prior ART | 1 (0.5) |
| Missing | 0 |
| When started ART | |
| Pre-HAART (before 1996) | 40 (18.2) |
| Early HAART (1996-2005) | 88 (40.0) |
| Modern HAART (since 2006) | 83 (37.7) |
| Missing | 9 (4.1) |
| Years since starting ART, Median (IQR) | 17 (9-21) |
| <10 | 53 (24.1) |
| 10-19 | 77 (35.0) |
| ≥20 | 81 (36.8) |
| Missing | 9 (4.1) |
| <20 | 130 (59.1) |
| ≥20 | 81 (36.8) |
| Missing | 9 (4.1) |
| Proportion of time since HIV diagnosis that participant was taking ART** | |

| | |
|---|---|
| >75% | 108 (49.1) |
| ≤75% | 99 (45.0) |
| Missing | 13 (5.9) |
| Stavudine and zidovudine exposure | |
| Stavudine | |
| Ever took | 77 (35.0) |
| Never took | 138 (62.7) |
| Missing | 5 (2.3) |
| Years took stavudine, median (IQR) | 2 (1-5) |
| Missing n (% of ever took) | 24 (28.6) |
| Zidovudine | |
| Ever took | 91 (41.4) |
| Never took | 124 (56.4) |
| Missing | 5 (2.3) |
| Years took zidovudine, median (IQR) | 3 (1-7) |
| Missing n (% of ever took) | 19 (20.9) |
| Ever took either stavudine or zidovudine | |
| Ever took either | 115 (52.3) |
| Never took either | 100 (45.5) |
| Missing | 5 (2.3) |

| | |
|---|---|
| Years took either stavudine or zidovudine***, | |
| median (IQR) | 5 (2-8) |
| Missing n (% of ever took) | 26 (22.6) |
| Current ART regimen | |
| Dolutegravir/abacavir/lamivudine | 27 (12.3) |
| Dolutegravir/emtricitabine/tenofovir alafenamide | 23 (10.5) |
| Elvitegravir/cobicistat/emtricitabine/tenofovir alafenamide | 22 (10.0) |
| Raltegravir/emtricitabine/tenofovir alafenamide | 16 (7.3) |
| Rilpivarine/emtricitabine/tenofovir alafenamide | 12 (5.5) |
| Darunavir/cobicistat/dolutegravir | 9 (4.1) |
| Nevirapine/emtricitabine/tenofovir alafenamide | 9 (4.1) |
| Efavirenz/emtricitabine/tenofovir disoproxil fumarate | 7 (3.2) |
| Dolutegravir/abacavir/lamivudine/maraviroc | 7 (3.2) |
| Nevirapine/abacavir/lamivudine | 5 (2.3) |
| Nevirapine/lamivudine/zidovudine | 4 (1.8) |
| Darunavir/ritonavir/raltegravir | 3 (1.4) |
| Raltegravir/emtricitabine/tenofovir disoproxil fumarate | 3 (1.4) |
| Other (≤ 2 participants per regimen) | 72 (32.7) |
| Not taking ART | 1 (0.5) |

*number of years from year of diagnosis to year of enrolment

**years on ART / years since diagnosis

***sum of the years on each individual drug

#### Supplementary Table S3. Characteristics of mutations associated with clonal haematopoiesis

| Characteristic | HIV negative N=51 n (%) | HIV positive N=81 n (%) | All N=132 n (%) |
|---|---|---|---|
| Variant allele fraction | | | |
| Median (IQR) | 4.0 (2.4 – 8.4) | 4.7 (2.6 – 8.6) | 4.6 (2.6 – 8.6) |
| ≤2 | 8 (15.7) | 7 (8.6) | 15 (11.4) |
| >2-≤5 | 20 (39.2) | 37 (45.7) | 57 (43.2) |
| >5-≤10 | 12 (23.5) | 18 (22.2) | 30 (22.7) |
| >10 | 11 (21.6) | 19 (23.5) | 30 (22.7) |
| Genes | | | |
| DNMT3A | 27 (53.0) | 37 (45.7) | 64 (48.5) |
| TET2 | 11 (21.6) | 16 (19.8) | 27 (20.5) |
| ASXL1 | 2 (3.9) | 13 (16.1) | 15 (11.4) |
| TP53 | 3 (5.9) | 4 (4.9) | 7 (5.3) |
| PPM1D | 2 (4.0) | 3 (3.7) | 5 (3.8) |
| JAK2 | 1 (2.0) | 2 (2.5) | 3 (2.3) |
| SF3B1 | 2 (4.0) | 1 (1.2) | 3 (2.3) |
| U2AF1 | 1 (2.0) | 2 (2.5) | 3 (2.3) |
| EZH2 | 1 (2.0) | 0 | 1 (0.8) |
| MPL | 0 | 1 (1.2) | 1 (0.8) |
| SETBP1 | 0 | 1 (1.2) | 1 (0.8) |
| STAG2 | 1 (2.0) | 0 | 1 (0.8) |

| | | | |
|---|---|---|---|
| STAT3 | 0 | 1 (1.2) | 1 (0.8) |
| Type of mutation | | | |
| Missense | 24 (47.1) | 29 (35.8) | 53 (40.2) |
| Frameshift | 18 (35.3) | 28 (34.6) | 46 (34.9) |
| Stop gained | 5 (9.8) | 16 (19.8) | 21 (15.9) |
| Splice donor | 2 (3.9) | 4 (4.9) | 6 (4.6) |
| Splice acceptor | 1 (2.0) | 2 (2.5) | 3 (2.3) |
| Inframe deletion | 1 (2.0) | 1 (1.2) | 2 (1.5) |
| Inframe insertion | 0 | 1 (1.2) | 1 (0.8) |
| Missense mutations | 24 (47.1) | 29 (35.8) | 53 (40.2) |
| Truncating[1] | 26 (51.0) | 50 (61.7) | 76 (57.6) |
| Inframe deletions or insertions | 1 (2.0) | 2 (2.5) | 3 (2.3) |
| Location of mutation within gene (DNMT3A only) | **N=27** | **N=37** | **N=64** |
| Mtase | 16 (59.3) | 19 (51.4) | 35 (54.7) |
| PWWP | 4 (14.8) | 7 (18.9) | 11 (17.2) |
| ADD | 5 (18.5) | 5 (13.5) | 10 (15.6) |
| Interaction with DNMT1 and DNMT3B | 2 (7.4) | 4 (10.8) | 6 (9.4) |
| Close to MTase | 0 | 2 (5.4) | 2 (3.1) |

[1]frame shift, stop gained, splice donor and splice acceptor mutations

#### Supplementary Table S4. Identified mutations associated with clonal haematopoiesis

| Subject | Gene | hgvsc | hgvsp | consequence | Variant allele frequency |
|---|---|---|---|---|---|
| 03-001 | TP53 | NM_000546.5:c.559+1G>A | p.? | splice_donor_variant | 2.04 |
| 03-001 | TP53 | NM_000546.5:c.481G>T | NP_000537.3:p.(Ala161Ser) | missense_variant | 2.85 |
| 03-006 | PPM1D | NM_003620.3:c.1434C>A | NP_003611.1:p.(Cys478*) | stop_gained | 7.23 |
| 03-008 | PPM1D | NM_003620.3:c.1442_1449del | NP_003611.1:p.(Ala481Valfs*5) | frameshift_variant | 11.75 |
| 03-009 | DNMT3A | NM_022552.4:c.2478+2T>C | p.? | splice_donor_variant | 13.04 |
| 03-013 | DNMT3A | NM_022552.4:c.1567G>T | NP_072046.2:p.(Glu523*) | stop_gained | 2.15 |
| 03-013 | DNMT3A | NM_022552.4:c.654dup | NP_072046.2:p.(Lys219Glnfs*34) | frameshift_variant | 2.77 |
| 03-015 | ASXL1 | NM_015338.5:c.1249C>T | NP_056153.2:p.(Arg417*) | stop_gained | 4.51 |
| 03-015 | ASXL1 | NM_015338.5:c.2485C>T | NP_056153.2:p.(Gln829*) | stop_gained | 16.20 |
| 03-020 | TET2 | NM_001127208.2:c.3734A>G | NP_001120680.1:p.(Tyr1245Cys) | missense_variant | 8.45 |
| 03-020 | TET2 | NM_001127208.2:c.4056del | NP_001120680.1:p.(Glu1352Aspfs*11) | frameshift_variant | 10.47 |
| 03-025 | DNMT3A | NM_022552.4:c.912del | NP_072046.2:p.(Trp305Glyfs*11) | frameshift_variant | 11.37 |
| 03-031 | TP53 | NM_000546.5:c.733G>C | NP_000537.3:p.(Gly245Arg) | missense_variant | 3.85 |
| 03-031 | EZH2 | NM_004456.4:c.2007C>G | NP_004447.2:p.(Ser669Arg) | missense_variant | 30.84 |
| 03-032 | DNMT3A | NM_022552.4:c.2207G>A | NP_072046.2:p.(Arg736His) | missense_variant | 2.73 |
| 03-032 | PPM1D | NM_003620.3:c.1568C>T | NP_003611.1:p.(Ala523Val) | missense_variant | 11.68 |
| 03-040 | TET2 | NM_001127208.2:c.1995_1998del | NP_001120680.1:p.(Asp666Ilefs*33) | frameshift_variant | 4.28 |
| 03-040 | DNMT3A | NM_022552.4:c.2206C>T | NP_072046.2:p.(Arg736Cys) | missense_variant | 6.34 |
| 03-045 | ASXL1 | NM_015338.5:c.2177del | NP_056153.2:p.(Lys726Argfs*18) | frameshift_variant | 2.77 |
| 03-049 | DNMT3A | NM_022552.4:c.1096C>A | NP_072046.2:p.(Arg366Ser) | frameshift_variant | 6.72 |
| 03-054 | JAK2 | NM_004972.3:c.1849G>T | NP_004963.1:p.(Val617Phe) | missense_variant | 1.93 |
| 03-054 | DNMT3A | NM_022552.4:c.820_821insTT | NP_072046.2:p.(Ala274Valfs*43) | frameshift_variant | 2.20 |
| 03-060 | DNMT3A | NM_022552.4:c.2183del | NP_072046.2:p.(Gly728Alafs*51) | frameshift_variant | 3.27 |
| 03-060 | DNMT3A | NM_022552.4:c.1717C>T | NP_072046.2:p.(Gln573*) | stop_gained | 6.97 |
| 03-064 | TET2 | NM_001127208.2:c.2083dup | NP_001120680.1:p.(Met695Asnfs*17) | frameshift_variant | 6.80 |
| 03-073 | TET2 | NM_001127208.2:c.4726_4733del | NP_001120680.1:p.(Val1576Leufs*35) | frameshift_variant | 4.70 |
| 03-077 | JAK2 | NM_004972.3:c.1849G>T | NP_004963.1:p.(Val617Phe) | missense_variant | 24.22 |

| | | | | | |
|---|---|---|---|---|---|
| 03-081 | DNMT3A | NM_022552.4:c.2253C>G | NP_072046.2:p.(Phe751Leu) | missense_variant | 13.98 |
| 03-084 | TET2 | NM_001127208.2:c.5602C>T | NP_001120680.1:p.(His1868Tyr) | missense_variant | 1.75 |
| 03-090 | DNMT3A | NM_022552.4:c.2577dup | NP_072046.2:p.(Trp860Metfs*4) | frameshift_variant | 3.52 |
| 03-090 | TET2 | NM_001127208.2:c.840dup | NP_001120680.1:p.(Asn281*) | frameshift_variant | 3.98 |
| 03-090 | DNMT3A | NM_022552.4:c.2206C>T | NP_072046.2:p.(Arg736Cys) | missense_variant | 5.52 |
| 03-091 | TET2 | NM_001127208.2:c.4997_5010del | NP_001120680.1:p.(Pro1666Glnfs*16) | frameshift_variant | 6.58 |
| 03-093 | DNMT3A | NM_022552.4:c.2644C>T | NP_072046.2:p.(Arg882Cys) | missense_variant | 7.39 |
| 03-096 | DNMT3A | NM_022552.4:c.1554+1del | p.? | splice_donor_variant | 2.20 |
| 03-096 | TET2 | NM_001127208.2:c.2474del | NP_001120680.1:p.(Ser825*) | frameshift_variant | 3.26 |
| 03-100 | TET2 | NM_001127208.2:c.5703_5704del | NP_001120680.1:p.(Phe1901Leufs*4) | frameshift_variant | 2.41 |
| 03-100 | DNMT3A | NM_022552.4:c.1883del | NP_072046.2:p.(Ala628Valfs*23) | frameshift_variant | 2.96 |
| 03-101 | DNMT3A | NM_022552.4:c.1583_1603dup | NP_072046.2:p.(Tyr528_Gln534dup) | inframe_insertion | 2.51 |
| 03-102 | DNMT3A | NM_022552.4:c.2676_2677del | NP_072046.2:p.(Trp893Glufs*27) | frameshift_variant | 2.07 |
| 03-112 | DNMT3A | NM_022552.4:c.2193_2195del | NP_072046.2:p.(Phe732del) | inframe_deletion | 2.66 |
| 03-127 | DNMT3A | NM_022552.4:c.2259del | NP_072046.2:p.(Trp753Cysfs*26) | frameshift_variant | 2.48 |
| 03-127 | DNMT3A | NM_022552.4:c.2409-1G>A | p.? | splice_acceptor_variant | 3.63 |
| 03-129 | DNMT3A | NM_022552.4:c.880G>T | NP_072046.2:p.(Glu294*) | stop_gained | 8.69 |
| 03-130 | TET2 | NM_001127208.2:c.3881A>G | NP_001120680.1:p.(Tyr1294Cys) | missense_variant | 15.18 |
| 03-131 | TET2 | NM_001127208.2:c.3680T>G | NP_001120680.1:p.(Val1227Gly) | missense_variant | 3.54 |
| 03-135 | DNMT3A | NM_022552.4:c.892G>T | NP_072046.2:p.(Gly298Trp) | missense_variant | 8.60 |
| 03-140 | DNMT3A | NM_022552.4:c.2645G>A | NP_072046.2:p.(Arg882His) | missense_variant | 4.88 |
| 03-143 | DNMT3A | NM_022552.4:c.2206C>T | NP_072046.2:p.(Arg736Cys) | missense_variant | 1.96 |
| 06-001 | MPL | NM_005373.2:c.1487C>T | NP_005364.1:p.(Thr496Ile) | missense_variant | 2.56 |
| 06-004 | TET2 | NM_001127208.2:c.4133G>A | NP_001120680.1:p.(Cys1378Tyr) | missense_variant | 5.00 |
| 06-006 | DNMT3A | NM_022552.4:c.1630_1631insT | NP_072046.2:p.(Arg544Leufs*2) | frameshift_variant | 3.41 |
| 06-006 | DNMT3A | NM_022552.4:c.886G>A | NP_072046.2:p.(Val296Met) | missense_variant | 3.64 |
| 06-007 | DNMT3A | NM_022552.4:c.2644C>T | NP_072046.2:p.(Arg882Cys) | missense_variant | 1.33 |
| 06-008 | DNMT3A | NM_022552.4:c.939G>A | NP_072046.2:p.(Trp313*) | stop_gained | 2.38 |
| 06-016 | DNMT3A | NM_022552.4:c.1816C>T | NP_072046.2:p.(Gln606*) | stop_gained | 20.96 |
| 06-018 | DNMT3A | NM_022552.4:c.2387del | NP_072046.2:p.(Gly796Valfs*6) | frameshift_variant | 5.83 |
| 06-021 | DNMT3A | NM_022552.4:c.942G>A | NP_072046.2:p.(Trp314*) | stop_gained | 4.33 |
| 06-026 | DNMT3A | NM_022552.4:c.2645G>A | NP_072046.2:p.(Arg882His) | missense_variant | 1.12 |

| | | | | | |
|---|---|---|---|---|---|
| 06-046 | DNMT3A | NM_022552.4:c.2645G>A | NP_072046.2:p.(Arg882His) | missense_variant | 1.50 |
| 06-050 | SF3B1 | NM_012433.2:c.2098A>G | NP_036565.2:p.(Lys700Glu) | missense_variant | 7.02 |
| 06-055 | DNMT3A | NM_022552.4:c.2645G>A | NP_072046.2:p.(Arg882His) | missense_variant | 1.11 |
| 06-062 | DNMT3A | NM_022552.4:c.2165del | NP_072046.2:p.(Gly722Alafs*57) | frameshift_variant | 8.03 |
| 06-067 | ASXL1 | NM_015338.5:c.4127dup | NP_056153.2:p.(Pro1377Serfs*3) | frameshift_variant | 5.30 |
| 06-072 | PPM1D | NM_003620.3:c.1642A>G | NP_003611.1:p.(Lys548Glu) | missense_variant | 2.39 |
| 06-072 | DNMT3A | NM_022552.4:c.1015-1G>A | p.? | splice_acceptor_variant | 7.63 |
| 10-002 | PPM1D | NM_003620.3:c.1423del | NP_003611.1:p.(Glu475Lysfs*8) | frameshift_variant | 4.22 |
| 10-003 | TET2 | NM_001127208.2:c.2079del | NP_001120680.1:p.(Lys693Asnfs*7) | frameshift_variant | 3.01 |
| 10-006 | DNMT3A | NM_022552.4:c.990G>A | NP_072046.2:p.(Trp330*) | stop_gained | 3.11 |
| 10-006 | DNMT3A | NM_022552.4:c.1066C>T | NP_072046.2:p.(Gln356*) | stop_gained | 11.79 |
| 10-011 | DNMT3A | NM_022552.4:c.2578T>C | NP_072046.2:p.(Trp860Arg) | missense_variant | 2.56 |
| 10-013 | DNMT3A | NM_022552.4:c.990G>A | NP_072046.2:p.(Trp330*) | stop_gained | 1.38 |
| 10-016 | DNMT3A | NM_022552.4:c.1652_1657delinsC | NP_072046.2:p.(Asn551Thrfs*25) | frameshift_variant | 7.09 |
| 10-024 | DNMT3A | NM_022552.4:c.2355_2356insCT | NP_072046.2:p.(Ser786Leufs*17) | frameshift_variant | 9.73 |
| 10-031 | DNMT3A | NM_022552.4:c.2206C>T | NP_072046.2:p.(Arg736Cys) | missense_variant | 1.71 |
| 10-034 | SF3B1 | NM_012433.2:c.1984C>T | NP_036565.2:p.(His662Tyr) | missense_variant | 1.47 |
| 10-034 | DNMT3A | NM_022552.4:c.2193C>G | NP_072046.2:p.(Phe731Leu) | missense_variant | 2.02 |
| 10-034 | TET2 | NM_001127208.2:c.244dup | NP_001120680.1:p.(Ser82Lysfs*2) | frameshift_variant | 2.73 |
| 10-036 | DNMT3A | NM_022552.4:c.2644C>T | NP_072046.2:p.(Arg882Cys) | missense_variant | 2.15 |
| 10-036 | ASXL1 | NM_015338.5:c.1900_1922del | NP_056153.2:p.(Glu635Argfs*15) | frameshift_variant | 12.64 |
| 10-063 | DNMT3A | NM_022552.4:c.1656del | NP_072046.2:p.(Asn552Lysfs*99) | frameshift_variant | 3.33 |
| 10-063 | DNMT3A | NM_022552.4:c.958C>T | NP_072046.2:p.(Arg320*) | stop_gained | 10.89 |
| 10-076 | TET2 | NM_001127208.2:c.5507_5508del | NP_001120680.1:p.(Val1836Glyfs*9) | frameshift_variant | 2.28 |
| 12-006 | ASXL1 | NM_015338.5:c.2035G>T | NP_056153.2:p.(Gly679*) | stop_gained | 7.05 |
| 12-008 | DNMT3A | NM_022552.4:c.1823del | NP_072046.2:p.(Phe608Serfs*43) | frameshift_variant | 3.51 |
| 18-013 | TP53 | NM_000546.5:c.772G>A | NP_000537.3:p.(Glu258Lys) | missense_variant | 8.38 |
| 18-013 | DNMT3A | NM_022552.4:c.846dup | NP_072046.2:p.(Glu283Argfs*41) | frameshift_variant | 12.95 |
| 18-014 | SF3B1 | NM_012433.2:c.2098A>G | NP_036565.2:p.(Lys700Glu) | missense_variant | 3.65 |
| 18-014 | DNMT3A | NM_022552.4:c.2479-1G>A | p.? | splice_acceptor_variant | 7.11 |
| 18-019 | DNMT3A | NM_022552.4:c.2193_2195del | NP_072046.2:p.(Phe732del) | inframe_deletion | 6.37 |
| 18-055 | ASXL1 | NM_015338.5:c.1900_1922del | NP_056153.2:p.(Glu635Argfs*15) | frameshift_variant | 20.31 |

| | | | | | |
|---|---|---|---|---|---|
| 20-003 | ASXL1 | NM_015338.5:c.3514del | NP_056153.2:p.(Ala1172Leufs*2) | frameshift_variant | 1.46 |
| 20-003 | DNMT3A | NM_022552.4:c.2311C>T | NP_072046.2:p.(Arg771*) | stop_gained | 2.73 |
| 20-006 | ASXL1 | NM_015338.5:c.2117del | NP_056153.2:p.(His706Leufs*19) | frameshift_variant | 5.03 |
| 20-006 | TET2 | NM_001127208.2:c.822del | NP_001120680.1:p.(Asn275Ilefs*18) | frameshift_variant | 5.34 |
| 20-006 | TET2 | NM_001127208.2:c.4133G>A | NP_001120680.1:p.(Cys1378Tyr) | missense_variant | 13.46 |
| 20-007 | STAT3 | NM_139276.2:c.1999G>T | NP_644805.1:p.(Val667Leu) | missense_variant | 4.61 |
| 20-008 | TET2 | NM_001127208.2:c.4567C>T | NP_001120680.1:p.(Gln1523*) | stop_gained | 4.68 |
| 20-012 | TET2 | NM_001127208.2:c.4399dup | NP_001120680.1:p.(Arg1467Lysfs*11) | frameshift_variant | 2.93 |
| 20-012 | DNMT3A | NM_022552.4:c.2207G>A | NP_072046.2:p.(Arg736His) | missense_variant | 3.38 |
| 20-018 | U2AF1 | NM_006758.2:c.470A>C | NP_006749.1:p.(Gln157Pro) | missense_variant | 3.78 |
| 20-025 | ASXL1 | NM_015338.5:c.2324del | NP_056153.2:p.(Leu775*) | frameshift_variant | 13.53 |
| 20-029 | TET2 | NM_001127208.2:c.2759del | NP_001120680.1:p.(Leu920*) | frameshift_variant | 2.18 |
| 21-001 | TET2 | NM_001127208.2:c.3733_3737del | NP_001120680.1:p.(Tyr1245Glyfs*21) | frameshift_variant | 13.14 |
| 21-004 | DNMT3A | NM_022552.4:c.1851+1G>A | p.? | splice_donor_variant | 2.21 |
| 21-009 | DNMT3A | NM_022552.4:c.2106T>A | NP_072046.2:p.(Asp702Glu) | missense_variant | 3.13 |
| 21-017 | ASXL1 | NM_015338.5:c.1247_1248del | NP_056153.2:p.(Leu416Profs*21) | frameshift_variant | 11.44 |
| 27-001 | DNMT3A | NM_022552.4:c.855+1G>A | p.? | splice_donor_variant | 1.31 |
| 27-002 | ASXL1 | NM_015338.5:c.2278C>T | NP_056153.2:p.(Gln760*) | stop_gained | 16.69 |
| 27-003 | STAG2 | NM_006603.4:c.192_193insGA | NP_006594.3:p.(Ser65Aspfs*7) | frameshift_variant | 18.38 |
| 27-008 | ASXL1 | NM_015338.5:c.2535dup | NP_056153.2:p.(Ser846Glnfs*5) | frameshift_variant | 7.13 |
| 27-008 | SETBP1 | NM_015559.2:c.2602G>A | NP_056374.2:p.(Asp868Asn) | missense_variant | 16.00 |
| 27-008 | U2AF1 | NM_006758.2:c.470A>C | NP_006749.1:p.(Gln157Pro) | missense_variant | 41.75 |
| 27-008 | TP53 | NM_000546.5:c.455C>T | NP_000537.3:p.(Pro152Leu) | missense_variant | 42.75 |
| 27-015 | ASXL1 | NM_015338.5:c.2143A>T | NP_056153.2:p.(Arg715*) | stop_gained | 16.25 |
| 27-016 | TET2 | NM_001127208.2:c.1669C>T | NP_001120680.1:p.(Gln557*) | stop_gained | 2.71 |
| 27-019 | TP53 | NM_000546.5:c.542G>A | NP_000537.3:p.(Arg181His) | missense_variant | 2.47 |
| 30-001 | TET2 | NM_001127208.2:c.3311_3312del | NP_001120680.1:p.(Phe1104Tyrfs*25) | frameshift_variant | 14.14 |
| 30-003 | DNMT3A | NM_022552.4:c.2645G>A | NP_072046.2:p.(Arg882His) | missense_variant | 2.15 |
| 30-009 | JAK2 | NM_004972.3:c.1849G>T | NP_004963.1:p.(Val617Phe) | missense_variant | 4.73 |
| 30-009 | TET2 | NM_001127208.2:c.2626C>T | NP_001120680.1:p.(Gln876*) | stop_gained | 5.01 |
| 30-014 | DNMT3A | NM_022552.4:c.2645G>A | NP_072046.2:p.(Arg882His) | missense_variant | 45.24 |
| 30-016 | DNMT3A | NM_022552.4:c.1809_1810del | NP_072046.2:p.(Arg604Alafs*7) | frameshift_variant | 1.95 |

| | | | | | |
|---|---|---|---|---|---|
| 30-017 | U2AF1 | NM_006758.2:c.470A>C | NP_006749.1:p.(Gln157Pro) | missense_variant | 6.56 |
| 30-019 | DNMT3A | NM_022552.4:c.2645G>A | NP_072046.2:p.(Arg882His) | missense_variant | 16.15 |
| 30-021 | TET2 | NM_001127208.2:c.4714C>T | NP_001120680.1:p.(Arg1572Trp) | missense_variant | 5.37 |
| 30-023 | DNMT3A | NM_022552.4:c.2640G>T | NP_072046.2:p.(Met880Ile) | missense_variant | 1.96 |
| 30-025 | DNMT3A | NM_022552.4:c.2645G>A | NP_072046.2:p.(Arg882His) | missense_variant | 2.54 |
| 30-027 | TP53 | NM_000546.5:c.659A>G | NP_000537.3:p.(Tyr220Cys) | missense_variant | 5.86 |
| 30-028 | DNMT3A | NM_022552.4:c.855+1G>A | p.? | splice_donor_variant | 1.71 |
| 30-028 | TET2 | NM_001127208.2:c.3781C>T | NP_001120680.1:p.(Arg1261Cys) | missense_variant | 8.57 |
| 30-028 | ASXL1 | NM_015338.5:c.1249C>T | NP_056153.2:p.(Arg417*) | stop_gained | 35.82 |

#### Supplementary Table S5. Sensitivity analysis of the association between HIV infection and the presence of having ≥1 CH mutation

**Supplementary Table S5.A. Mutations with variant allele fraction>2% only**[1]: 88 (19.7%) of 446 participants (200 HIV positive, 226 HIV negative) had ≥1 CH mutation with VAF>2%

| | No CH mutation N=358 | ≥1 CH mutation N=88 | OR (95% CI) Unadjusted | p-value unadjusted | OR (95% CI) adjusted for age, gender, smoking and sexual orientation | p-value adjusted for age, gender, smoking and sexual orientation |
|---|---|---|---|---|---|---|
| HIV status | | | | | | |
| HIV negative | 195 (54.5) | 31 (35.2) | Ref | | | |
| HIV positive | 163 (45.5) | 57 (64.8) | 2.20 (1.36-3.57) | 0.001 | 2.47 (1.48-4.11) | <0.001 |
| Missing | 0 | 0 | | | | |
| Age | | | | | | |
| Median age (IQR) | 63 (59-68) | 66 (61-71) | 1.07 (1.04-1.12) | <0.001 | 1.08 (1.04-1.13) | <0.001 |
| Missing | 0 | 0 | | | | |
| Gender | | | | | | |
| Female | 13 (3.6) | 4 (4.6) | Ref | | | |
| Male | 345 (96.4) | 84 (95.5) | 0.79 (0.25-2.49) | 0.69 | 0.53 (0.11-2.49) | 0.42 |
| Missing | 0 | 0 | | | | |
| Sexual orientation | | | | | | |

| | | | | | | |
|---|---|---|---|---|---|---|
| Other | 35 (9.8) | 8 (9.1) | Ref | | | |
| MSM | 313 (87.4) | 77 (87.5) | 1.08 (0.48-2.41) | 0.86 | 1.30 (0.45-3.77) | 0.63 |
| Missing | 10 (2.8) | 3 (3.4) | | | | |
| Smoking | | | | | | |
| Never smoked | 162 (45.3) | 33 (37.5) | Ref | | | |
| Ever smoked | 154 (43.0) | 46 (52.3) | 1.47 (0.89-2.41) | 0.13 | 1.39 (0.83-2.33) | 0.21 |
| Missing | 42 (11.7) | 9 (10.2) | | | | |

[1]In this model, if a participant had multiple mutations, they were classified as having any CH mutation if any of the mutations had a VAF >2.

**Supplementary Table S5.B. Mutations in DNMT3A, TET2 and ASXL1 only**[1]: 85 (19.1%) of 446 participants (200 HIV positive, 226 HIV negative) had ≥1 CH mutation in DNMT3A, TET2 or ASXL1

| | No CH mutation (N=361) | ≥1 CH mutation (N=85) | OR (95% CI) Unadjusted | p-value unadjusted | OR (95% CI) adjusted for age, gender, smoking and sexual orientation | p-value adjusted for age, gender, smoking and sexual orientation |
|---|---|---|---|---|---|---|
| HIV status | | | | | | |
| HIV negative | 194 (53.7) | 32 (37.7) | Ref | | | |
| HIV positive | 167 (46.3) | 53 (62.4) | 1.92 (1.18-3.13) | 0.008 | 2.08 (1.26-3.46) | 0.004 |
| Missing | 0 | 0 | | | | |
| Age | | | | | | |
| Median age (IQR) | 63 (59-68) | 66 (61-70) | 1.07 (1.03-1.11) | <0.001 | 1.07 (1.03-1.12) | <0.001 |
| Missing | 0 | 0 | | | | |
| Gender | | | | | | |
| Female | 13 (3.6) | 4 (4.7) | Ref | | | |
| Male | 348 (96.4) | 81 (95.3) | 0.76 (0.24-2.38) | 0.63 | 0.64 (0.14-2.95) | 0.57 |
| Missing | 0 | 0 | | | | |
| Sexual orientation | | | | | | |
| Other | 34 (9.4) | 9 (10.6) | Ref | | | |
| MSM | 315 (87.3) | 75 (88.2) | 0.90 (0.41-1.96) | 0.79 | 1.02 (0.37-2.80) | 0.97 |

| | | | | | | |
|---|---|---|---|---|---|---|
| Missing | 12 (3.3) | 1 (1.2) | | | | |
| Smoking | | | | | | |
| Never smoked | 163 (45.2) | 32 (37.7) | Ref | | | |
| Ever smoked | 156 (43.2) | 44 (51.8) | 1.44 (0.87-2.38) | 0.16 | 1.39 (0.82-2.33) | 0.22 |
| Missing | 42 (11.6) | 9 (10.6) | | | | |

[1]In this model, if a participant had multiple mutations, they were classified as having any CH mutation if any of the mutations were in DNMT3A, TET2 or ASXL1.

**Supplementary Table S5.C. Men only**: N=95 (22.1%) of 429 male participants (216 HIV positive, 213 HIV negative) had ≥1 CH mutation

| | No CH mutation (N=334) | ≥1 CH mutation (N=95) | OR (95% CI) Unadjusted | p-value unadjusted | OR (95% CI) adjusted for age, smoking and sexual orientation | p-value adjusted for age, smoking and sexual orientation |
|---|---|---|---|---|---|---|
| HIV status | | | | | | |
| HIV negative | 117 (53.0) | 36 (37.9) | Ref | | | |
| HIV positive | 157 (47.0) | 59 (62.1) | 1.85 (1.16-2.95) | 0.01 | 2.04 (1.26-3.32) | 0.004 |
| Missing | 0 | 0 | | | | |
| Age | | | | | | |
| Median age (IQR) | 63 (59-68) | 66 (61-71) | 1.08 (1.04-1.12) | <0.001 | 1.08 (1.04-1.13) | <0.001 |
| Missing | 0 | 0 | | | | |
| Sexual orientation | | | | | | |
| Other | 21 (6.3) | 6 (6.3) | Ref | | | |
| MSM | 304 (91.0) | 86 (90.5) | 0.99 (0.39-2.53) | 0.98 | 0.99 (0.38-2.58) | 0.98 |
| Missing | 9 (2.7) | 3 (3.2) | | | | |
| Smoking | | | | | | |
| Never smoked | 152 (45.5) | 37 (39.0) | Ref | | | |
| Ever smoked | 143 (42.8) | 47 (49.5) | 1.35 (0.83-2.20) | 0.23 | 1.27 (0.77-2.10) | 0.36 |
| Missing | 39 (11.7) | 11 (11.6) | | | | |

**Supplementary Table S5.D. MSM only**: 86 (22.1%) of 390 MSM participants (201 HIV positive, 189 HIV negative) had ≥1 CH mutation

| | **No CH mutation (N=304)** | **≥1 CH mutation (N=86)** | **OR (95% CI) Unadjusted** | **p-value unadjusted** | **OR (95% CI) adjusted for age and smoking** | **p-value adjusted for age and smoking** |
|---|---|---|---|---|---|---|
| HIV status | | | | | | |
| HIV negative | 157 (51.6) | 32 (37.2) | Ref | | | |
| HIV positive | 147 (48.4) | 54 (62.8) | 1.80 (1.10-2.95) | 0.02 | 1.98 (1.19-3.31) | 0.009 |
| Missing | 0 | 0 | | | | |
| Age | | | | | | |
| Median age (IQR) | 63 (58-67) | 66 (61-71) | 1.08 (1.03-1.12) | <0.001 | 1.08 (1.04-1.13) | <0.001 |
| Missing | 0 | 0 | | | | |
| Smoking | | | | | | |
| Never smoked | 141 (46.4) | 34 (39.5) | Ref | | | |
| Ever smoked | 129 (42.4) | 41 (47.7) | 1.32 (0.79-2.20) | 0.29 | 1.24 (0.73-2.10) | 0.43 |
| Missing | 34 (11.2) | 11 (12.8) | | | | |

**Supplementary Table S6. Univariate analysis of risk factors for having ≥ one CH mutation among HIV positive participants (N=220)**

| | No CH Mutation N=159 | Any CH Mutation N=61 | Univariate OR (95% CI) | Univar p-value |
|---|---|---|---|---|
| Median age (IQR) | 62 (59-67) | 66 (61-71) | 1.11 (1.05-1.17) | <0.001 |
| Missing | 0 | 0 | | |
| Gender | | | | |
| Female | 2 (1.3) | 2 (3.3) | Ref | |
| Male | 157 (98.7) | 59 (96.7) | 0.38 (0.05-2.73) | 0.33 |
| Missing | 0 | 0 | | |
| Sexual orientation | | | | |
| Other | 9 (5.7) | 6 (9.8) | Ref | |
| MSM | 147 (92.5) | 54 (88.5) | 0.55 (0.19-1.62) | 0.28 |
| Missing | 3 (1.9) | 1 (1.6) | | |
| Years since HIV diagnosis, Median (IQR)* | 23 (14-30) | 26 (16-31) | 1.01 (0.98-1.04) | 0.69 |
| <10 | 14 (8.8) | 7 (11.5) | Ref | |
| 10-19 | 48 (30.2) | 16 (26.2) | 0.67 (0.23-1.94) | 0.46 |
| 20-29 | 47 (29.6) | 18 (29.5) | 0.77 (0.27-2.21) | 0.62 |
| ≥30 | 46 (28.9) | 20 (32.8) | 0.87 (0.31-2.48) | 0.79 |
| Missing | 4 (2.5) | 0 | | |

| | | | | |
|---|---|---|---|---|
| <20 | 62 (39.0) | 23 (37.7) | Ref | |
| ≥20 | 93 (58.5) | 38 (62.3) | 1.10 (0.60-2.03) | 0.76 |
| Missing | 4 (2.5) | 0 | | |
| CD4 nadir, Median (IQR) | 242 (133-372) | 271 (170-400) | 1.00 (0.99-1.00) | 0.26 |
| ≥500 | 20 (12.6) | 8 (13.1) | Ref | |
| 200-499 | 80 (50.3) | 34 (55.7) | 1.06 (0.43-2.65) | 0.90 |
| <200 | 56 (35.2) | 19 (31.2) | 0.85 (0.32-2.24) | 0.74 |
| Missing | 3 (1.9) | 0 | | |
| ≥200 | 100 (62.9) | 42 (68.9) | Ref | |
| <200 | 56 (35.2) | 19 (31.2) | 0.81 (0.43-1.52) | 0.51 |
| Missing | 3 (1.9) | 0 | | |
| Current CD4 count | | | | |
| Median (IQR) | 650 (483-865) | 626 (444-783) | 1.00 (0.99-1.00) | 0.62 |
| ≥500 | 115 (72.3) | 38 (62.3) | Ref | |
| 350-499 | 24 (15.1) | 15 (24.6) | 1.89 (0.90-3.97) | 0.09 |
| <350 | 18 (11.3) | 8 (13.1) | 1.35 (0.54-3.34) | 0.52 |
| Missing | 2 (1.3) | 0 | | |
| ≥500 | 115 (72.3) | 38 (62.3) | Ref | |
| <500 | 42 (26.4) | 23 (37.7) | 1.66 (0.89-3.10) | 0.11 |
| Missing | 2 (1.3) | 0 | | |
| Current HIV viral load | | | | |

| | | | | |
|---|---|---|---|---|
| <40 | 152 (95.6) | 58 (95.1) | Ref | |
| ≥40 | 7 (4.4) | 3 (4.9) | 1.12 (0.28-4.49) | 0.87 |
| Missing | 0 | 0 | | |
| When started ART | 129 (81.1) | 42 (68.9) | | |
| Modern HAART (since 2006) | 62 (39.0) | 21 (34.4) | Ref | |
| Early HAART (1996-2005) | 67 (42.1) | 21 (34.4) | 0.93 (0.46-1.86) | 0.83 |
| Pre-HAART (before 1996) | 26 (16.4) | 14 (23.0) | 1.59 (0.70-3.60) | 0.27 |
| Missing | 4 (2.5) | 5 (8.2) | | |
| Years from diagnosis to start of ART, Median (IQR) | 5 (1-11) | 4.5 (1-10) | 0.99 (0.94-1.04) | 0.66 |
| Within 1 year | 36 (22.6) | 12 (19.7) | Ref | |
| 1-4 years | 35 (22.0) | 16 (26.2) | 1.37 (0.57-3.31) | 0.48 |
| 5-9 years | 32 (20.1) | 13 (21.3) | 1.22 (0.49-3.05) | 0.67 |
| ≥10 years | 48 (30.2) | 15 (24.6) | 0.94 (0.29-2.25) | 0.88 |
| Missing | 8 (5.0) | 5 (8.2) | | |
| Years of HIV therapy, Median (IQR) | 17 (10-21) | 19 (9-23) | 1.01 (0.97-1.06) | 0.51 |
| <10 | 38 (23.9) | 15 (25.6) | Ref | |
| 10-19 | 62 (39.0) | 15 (24.6) | 0.61 (0.27-1.39) | 0.24 |
| ≥20 | 53 (33.3) | 24 (39.3) | 1.20 (0.56-2.56) | 0.64 |
| Missing | 4 (2.5) | 5 (8.2) | | |
| <20 | 100 (62.9) | 30 (49.2) | Ref | |

| | | | | |
|---|---|---|---|---|
| ≥20 | 55 (34.6) | 26 (42.6) | 1.58 (0.85-2.93) | 0.15 |
| Missing | 4 (2.5) | 5 (8.2) | | |
| Proportion of time since HIV diagnosis that participant was taking ART** | | | | |
| >75% | 79 (49.7) | 29 (47.5) | Ref | |
| ≤75% | 72 (45.3) | 27 (44.3) | 1.02 (0.55-1.89) | 0.95 |
| Missing | 8 (5.0) | 5 (8.2) | | |
| Ever took stavudine | | | | |
| No | 103 (64.8) | 35 (57.4) | Ref | |
| Yes | 54 (34.0) | 23 (37.7) | 1.25 (0.67-2.33) | 0.48 |
| Missing | 2 (1.3) | 3 (4.9) | | |
| Ever took zidovudine | | | | |
| No | 90 (56.6) | 34 (55.7) | Ref | |
| Yes | 67 (42.1) | 24 (39.3) | 3.97 (0.64-24.81) | 0.86 |
| Missing | 2 (1.3) | 3 (4.9) | | |
| Ever took stavudine or zidovudine | | | | |
| No | 74 (46.5) | 26 (42.6) | Ref | |
| Yes | 83 (52.2) | 32 (52.5) | 1.10 (0.60-2.01) | 0.76 |
| Missing | 2 (1.3) | 3 (4.9) | | |
| Current or prior HBV or HCV infection | | | | |
| No | 132 (83.0) | 51 (83.6) | Ref | |
| Yes | 27 (17.0) | 10 (16.4) | 0.96 (0.43-2.12) | 0.92 |

| | | |
|---|---|---|
| Missing | 0 | 0 |

*number of years from year of diagnosis to year of enrolment

**years on ART / years since diagnosis

**Supplementary Figure S1: Variant allele fraction by duration of HIV, among HIV positive participants[1]**

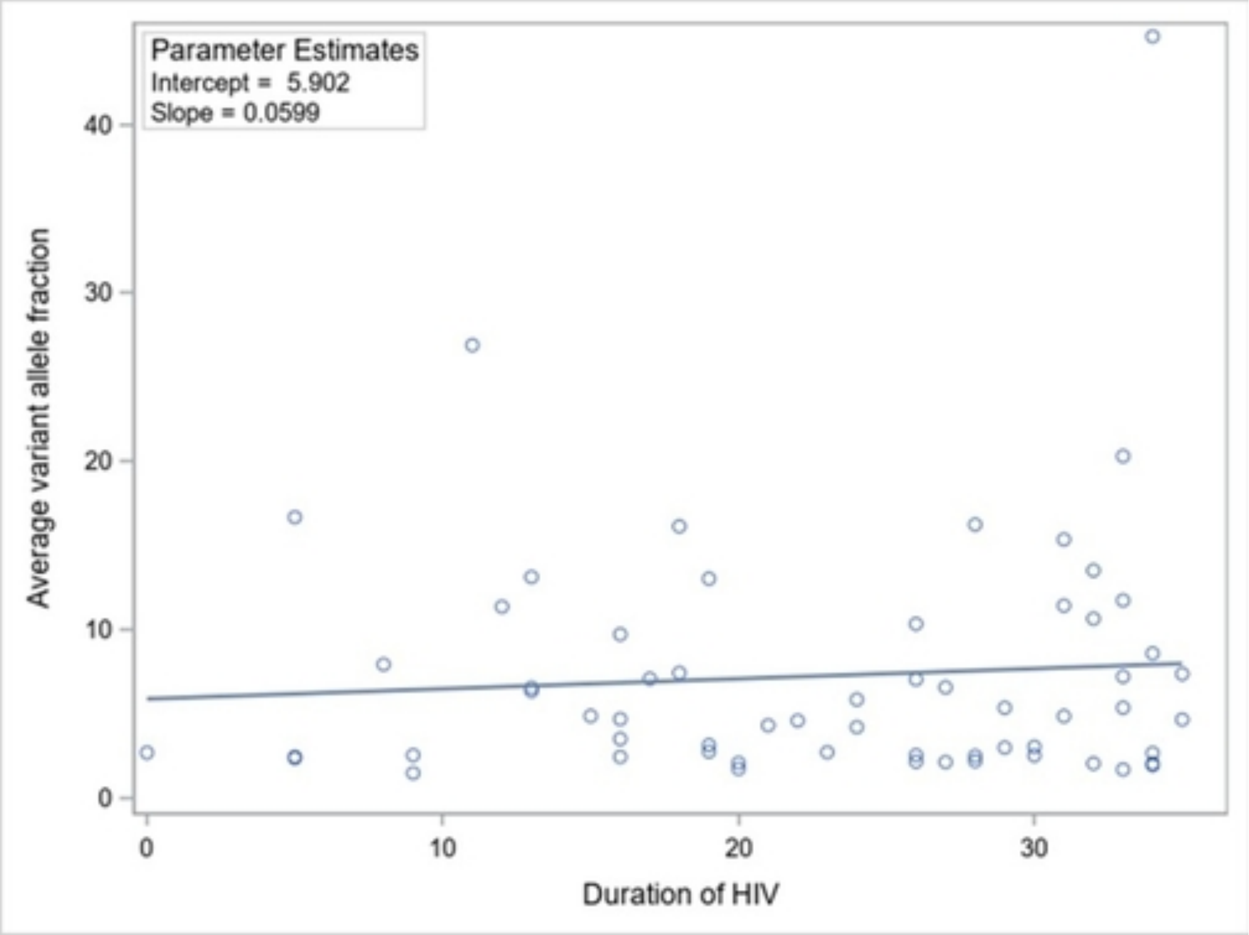

[1]Mean average variant allele fraction was calculated if there was more than one CH mutation present

**Supplementary Figure S2. Box plots of the distribution of blood cell characteristics and inflammatory markers.**

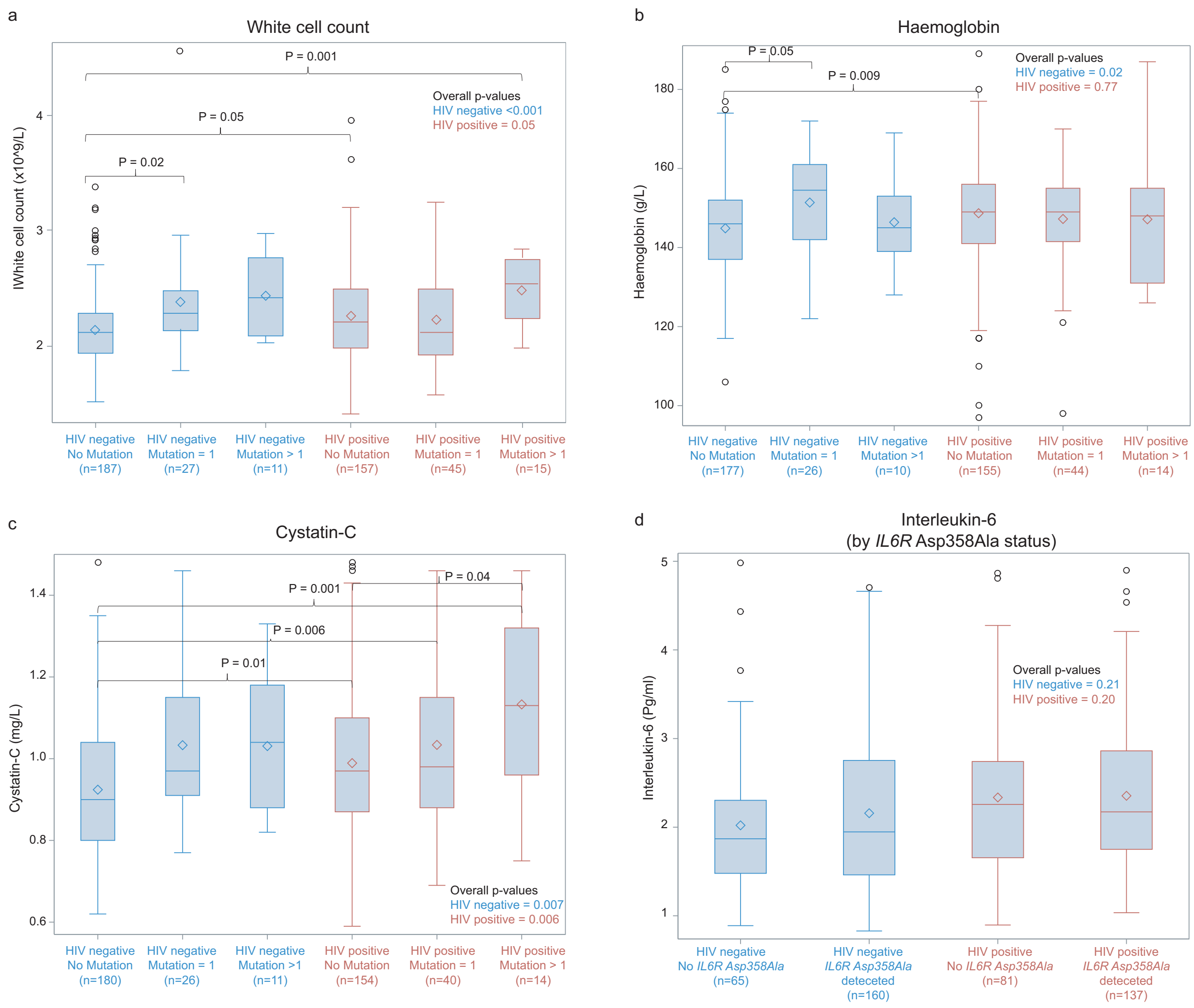

Overall p-values (figure inset) were generated using the Kruskal-Wallis test. P-values across specific groups (brackets) were generated using post-hoc pairwise two-sided multiple comparison analysis using the Dwass, Steel, Critchlow-Fligner Method.

Panel A. White cell count by HIV status and presence of 0, 1 and >1 CH mutation

Panel B. Haemoglobin by HIV status and presence of 0, 1 and >1 CH mutation. Haemoglobin was assessed among male participants only due to physiologic differences in haemoglobin across gender, and low numbers of women in our study.

Panel C. Cystatin C* by HIV status and presence of 0, 1 and >1 CH mutation

Panel D. Interleukin-6* by *IL6R p. Asp358Ala* status and HIV status

*Outliers have been removed for box plot presentation only; p-values were calculated including outliers. Outliers were defined as values greater than 1.5 times the IQR above the third quartile.

**References**

1.Doig KD, Fellowes A, Bell AH, et al. PathOS: a decision support system for reporting high throughput sequencing of cancers in clinical diagnostic laboratories. Genome medicine 2017;9:38.

2.Richards S, Aziz N, Bale S, et al. Standards and guidelines for the interpretation of sequence variants: a joint consensus recommendation of the American College of Medical Genetics and Genomics and the Association for Molecular Pathology. Genetics in medicine : official journal of the American College of Medical Genetics 2015;17:405-24.
