## Supplementary material for "Age-related clonal haematopoiesis is more prevalent in older adults with HIV: the ARCHIVE study": ARCHIVE study protocol

### Age-related clonal haematopoiesis in an HIV evaluation cohort (ARCHIVE): a follow-on study of the Australian Positive & Peers Longevity Evaluation Study

#### Short Title: ARCHIVE

#### Sponsor: Kirby Institute, UNSW Sydney

PROJECT TEAM ROLES & RESPONSIBILITIES

Co-Principal Investigator: Dr Nila Dharan BA MD

Signature: Date:

Organisation: The Kirby Institute, UNSW Sydney

Department: Biostatistics and Databases Program

Position: Research Fellow

Responsibilities: Dr Dharan will be responsible for assisting with the overall conduct of the study including coordination with participating sites, logistics of samples for testing, and submission of the study to the HREC and communicating with the HREC, and interpretation of results.

Co-Principal Investigator: Dr Mark Polizzotto MB BS BMedSc PhD FRACP FRCPA

Signature: Date:

Organisation: The Kirby Institute, UNSW Sydney

Department: Therapeutic and Vaccine Research Program

Position: Program Head (Acting)

Responsibilities: Dr Polizzotto will be responsible for assisting with the overall conduct of the study including coordination of samples for testing and interpretation of results.

Co-Principal Investigator: Dr Kathy Petoumenos BSc MPH PhD

Signature: Date:

Organisation: The Kirby Institute, UNSW Sydney

Department: Biostatistics and Databases Program

Position: Associate Professor

Responsibilities: Dr Petoumenos will be responsible for assisting with the conduct of the study including communication and coordination with sites and interpretation of results.

**Summary**

| | |
|---|---|
| **Study Title** | Age-related clonal haematopoiesis in an HIV evaluation cohort (ARCHIVE): a follow-on study of the Australian Positive & Peers Longevity Evaluation Study |
| **Objectives** | Primary: Establish a longitudinal cohort of older people with and without HIV for the evaluation of genomic and other factors associated with aging<br>Secondary:<br>1. Measure the prevalence of somatic mutations associated with clonal haematopoiesis (CH) in adults with HIV over 55 years of age and compare with the prevalence in age-matched controls (without HIV).<br>a. Determine whether there is a correlation between the prevalence of somatic mutations and: a) elevated inflammatory markers; or b) the presence of comorbidities<br>b. Compare the results of testing plasma samples for mutations associated with CH with that of testing whole blood<br>2. Link data obtained in this study with that from national and state-wide registries to provide outcomes data for our cohort such as hospitalizations, deaths, cancer diagnoses, and data from the Pharmaceutical Benefits Scheme and the Medicare Benefits Schedule |
| **Study design** | The study design is a longitudinal cohort study with a single initial proposed evaluation and potential follow-up visits as described in Study Duration below. |
| **Planned sample size** | 440 persons will be recruited in total; 220 participants with HIV and 220 participants without HIV. |
| **Selection criteria** | • Men and women aged >55 years<br>• For participants without HIV: an HIV negative test within 12 months prior to enrolment. If no HIV-negative test result is available, then potential participants will be tested for HIV as part of standard of care, if indicated (see details in section 4.1 below).<br>• Willing and able to provide consent to participate in a longitudinal cohort study including 1) consent to providing samples for genomics analysis 2) consent to linking their data to national and state-wide data registries; and 3) consent to participate in future follow-up studies as outlined. |
| **Study procedures** | There is one study visit for the proposed initial evaluation. During this study visit, one blood sample will be collected, participants will complete an online questionnaire, and personnel at the clinical sites will be asked to record information from the participant's chart.<br>Participants will be asked to fill out an on-line survey where they will be asked about health and lifestyle questions such as current or previous diagnoses of medical conditions (cardiovascular, haematologic, pulmonary, infectious, renal, endocrine, musculoskeletal, neurologic and psychiatric conditions), smoking and drug use, and history of fractures and/or falls. In addition, we will ask questions regarding antiretroviral (ART) use (for participants with HIV). For participants without HIV, we will ask whether they have ever taken PrEP, and if so, which medication(s) and dates they took it.<br>Sites will review the participant's chart and abstract information on the patient's medical history including diagnoses of medical conditions, the occurrence of medical events and use of ART.<br>The clinical samples obtained at the first study visit will include one blood |

| | |
|---|---|
| | sample that will be tested for mutations associated with clonal haematopoiesis, a complete blood count and inflammatory markers. |
| **Statistical considerations** | Sample size: We will recruit 440 participants: 220 with HIV and 220 without HIV. We will assume that 10% of our participants without HIV will have somatic mutations associated with clonal haematopoiesis and will power our study to detect an increase in prevalence (doubling) from 10% to 20% prevalence among participants with HIV. Testing 220 participants without HIV and 220 participants with HIV will give us 80% power to detect an increase to 20% prevalence of mutations among participants with HIV compared to the background mutation rate of 10% among participants without HIV.<br><br>We will also test all participants' plasma samples for the presence of these mutations and compare the results to those found from testing whole blood. The purpose of this comparison is to validate using plasma samples for detection of somatic mutations. Here we will perform correlation analysis comparing mutant allele fraction of whole blood with plasma using the spearman method. Our evaluation of an association between somatic mutations and inflammatory markers and clinical outcomes (comorbidities and medical events) will be exploratory.<br><br>Analysis plan: The prevalence of somatic mutations associated with clonal haematopoiesis will be summarized by HIV infection status. Conventional statistics will be used to compare the results across the two groups and determine if there are any statistically significant differences. For the plasma validation analysis, results of genomic testing of whole blood samples will be compared to those of plasma in terms of the number and type of mutation determined in each paired sample.<br><br>Health information collected for this study may also be linked to other state and national health registries, cancer registries and hospitalization data (i.e. admitted patients). This linkage will depend on the requirements of this and future research projects |
| **Study duration** | 5 years. First follow-up visit will be scheduled 6-12 months after the first visit, and subsequent visits will be scheduled every 1-2 years (maximum of 6 study visits over five years). The timing of subsequent visits will be determined by the results of this initial evaluation. |

TABLE OF CONTENTS

1. BACKGROUND 6
1.1. DISEASE BACKGROUND 6
1.2. RATIONALE FOR PERFORMING THE STUDY 6
2. STUDY OBJECTIVES 7
2.1. PRIMARY OBJECTIVE 7
2.2. SECONDARY OBJECTIVES 7
3. STUDY DESIGN 8
3.1. DESIGN 8
3.2. STUDY GROUPS 8
3.3. NUMBER OF PARTICIPANTS 8
3.4. NUMBER OF SITES 8
3.5. DURATION 8
4. PARTICIPANT SECTION 8
4.1. INCLUSION CRITERIA 8
4.2. EXCLUSION CRITERIA 9
5. STUDY OUTLINE 9
5.1. INVESTIGATION PLAN 9
5.2. STUDY PROCEDURE RISKS 11
5.3. RECRUITMENT AND SCREENING 11
5.4. INFORMED CONSENT PROCESS 11
5.5. ENROLMENT PROCEDURE 12
6. TISSUE COLLECTION/BIOBANKING 12
7. SAFETY 12
7.1. EARLY TERMINATION 12
8. OUTCOMES AND FUTURE PLANS 13
9. STATISTICAL CONSIDERATIONS 13
10. CONFIDENTIALITY AND STORAGE AND ARCHIVING OF STUDY DOCUMENTS 14
11. OTHER STUDY DOCUMENTS 14
12. RESOURCES 14
13. REFERENCES 15

#### 1. BACKGROUND

##### 1.1. Disease Background

Despite effective antiretroviral therapy (ART), non-AIDS related comorbidities, especially cardiovascular[1-3] and non-AIDS defining malignancies[3], occur in persons with HIV in greater number[1,2] and at an earlier age[4,5] than in people without HIV. This phenomenon is found even among people with HIV who have high rates of viral suppression on ART. The mechanism for developing these comorbidities is likely multifactorial[6], including inflammation, immunodeficiency, hypercoagulability, direct viral effects[7], and lifestyle factors[8-10]. With highly effective and easily tolerated modern ART regimens, people with HIV are living longer[5,11] and developing comorbidities common to older persons such as cancer and heart disease. Optimal prevention, detection and management of comorbidities are critical for effective long-term care of people with HIV.

Interest in studying the genetic determinants of aging and the development of comorbidities in the general population has increased in recent years. For example, new studies in the general population have shown that clonal haematopoiesis (CH), the expansion of hematopoietic cell lines driven by certain somatic mutations in a subset of key genes, increases with age among patients *without* hematologic malignancies[12,13]. These mutations are mainly associated with the development of myelodysplastic syndrome, which is a haematological malignancy associated with increased risk of developing acute myeloid leukaemia. Interestingly, the somatic mutations associated with CH have found to be associated not only with hematologic malignancies but also with all-cause mortality and cardiovascular events. To our knowledge, studies evaluating people with HIV for the presence of somatic mutations associated with CH have not been performed.

To effectively study genomic factors that influence the development of comorbidities among people with HIV, longitudinal cohorts must be established for periodic follow-up data collection over time. The Australian Positive and Peers Longevity Evaluation Study (APPLES) was a cohort study of 446 men who have sex with men (MSM) $\geq$ 55 years, both with and without HIV. Data collected on participants included clinical information, a self-completed health and lifestyle survey, and a blood sample collection for biomarker assessment and storage. In total, 228 men with HIV and 218 men without HIV were recruited, with a median age of 63 years (IQR: 59-67). The study proposed here will evaluate for genomic factors associated with aging and the development of comorbidities, including mutations associated with CH, among persons who previously participated in APPLES and newly-recruited men and women with and without HIV over age 55 that did not take part in APPLES.

The APPLES site network provides a unique already-established network that can be used to easily recruit participants to this evaluation of CH. Leveraging this existing group of clinical sites, we will establish a longitudinal cohort of older individuals with and without HIV in order to evaluate genomic factors associated with aging and the development of comorbidities. Our first focus will be to evaluate mutations associated with CH to answer the following questions: 1) What is the rate of somatic mutations associated with CH in people with HIV and how does it compare to the rates of these mutations in age-matched controls (people without HIV); 2) Is there an association between the presence of these mutations and the development of comorbidities and/or elevated inflammatory markers; and 3) Can plasma samples be used to detect mutations associated with CH. In this last aim, we will attempt to validate our detection of mutations in whole blood (the standard sample that is usually collected for genomics studies) in paired plasma samples. Considering that a number of HIV cohort studies have collected and stored plasma samples from older adults, whereas few have collected samples specifically for genomic analysis, validating the use of plasma samples for these types of genomics analyses would open up many more study populations for testing. In the APPLES cohort, retrospective genomics testing on stored samples would be consistent with the consent for initial participation in the study.

##### 1.2. Rationale for Performing the Study

The scientific precedent for the link between viruses and malignancies has been established in several clinical settings. The Human Papilloma Virus is a known risk factor for the development of anal and cervical dysplasia malignancies, and the Epstein Barr Virus is a known risk factor for the development of lymphoma. As above, somatic mutations associated with CH have been linked to the development of haematologic malignancies as well as cardiovascular and other outcomes including all-cause mortality in persons without HIV. In this study, we seek to evaluate for this link in HIV patients, and compare to a similar cohort of persons without HIV for comparison. The increased incidence of cardiovascular disease and malignancies in the HIV population, even despite optimal antiretroviral therapy, has not been explained. Given this, the clear link between chronic viral infections and the development of malignancies, and the established link between CH and cardiovascular and malignant outcomes in the general population, evaluating for CH among persons with HIV is of paramount importance.

Our findings will impact scientific knowledge about underlying mechanisms that drive comorbid conditions in the HIV population and lead to future investigations designed to: 1) further define the mechanisms behind the development of these somatic mutations as related to HIV infection such as viral effects, inflammation or other; 2) identify prominent mutations or combinations of mutations that may be associated with the greatest risk of cancer or other events, with the possible future application of routine screening in at-risk populations; 3) evaluate measures of prevention targeted to reduce the development of these mutations and therefore the progression to cancer or the development of other non-AIDS related comorbidities, adverse outcomes and events in the aging HIV population; and 4) evaluate for possible therapies targeting these somatic mutations such as emerging epigenetic therapies.

As above, the APPLES site network provides a unique established network that can be used to easily recruit participants to this evaluation. The involved sites have access to the necessary patient populations (older adults both with and without HIV) and are already experienced in enrolling subjects into studies involving data collection from self-report, chart review and clinical sample collection. In addition, the cohorts are well-matched for age and lifestyle factors; therefore, any differences in CH found between the two groups are less likely to be related to lifestyle factors, supporting our hypothesis that HIV infection leads to the development of CH. Lastly, retrospective genomics testing on stored samples would be consistent with the consent for initial participation in the study, so the original cohort will be more likely to be open to the idea of participating in a genomics study.

#### 2. STUDY OBJECTIVES

We hypothesize that individuals with HIV will have a higher age-specific prevalence of mutations associated with clonal haematopoiesis than the general population, and that these mutations may be associated with HIV viremia, elevated inflammatory markers and/or the presence of comorbidities and clinical events. To test this hypothesis our primary objective is to establish a longitudinal cohort of aging people with HIV and an age-matched cohort of people without HIV.

##### 2.1. PRIMARY OBJECTIVE

Establish a longitudinal cohort of people with and without HIV for the evaluation of genomic and other factors associated with aging

##### 2.2. SECONDARY OBJECTIVES

1. Measure the prevalence of somatic mutations associated with CH in adults with HIV over 55 years of age and compare with the prevalence in age-matched controls (people without HIV).
    a. Determine whether there is a correlation between the prevalence of somatic mutations and: a) elevated inflammatory markers; or b) the presence of comorbidities

b. Compare the results of testing plasma samples for mutations associated with CH with that of testing whole blood
2. Link data obtained in this study with that from national and state-wide registries to provide outcomes data for our cohort such as hospitalizations, deaths, cancer diagnoses, and data from the Pharmaceutical Benefits Scheme and the Medicare Benefits Schedule

#### 3. STUDY DESIGN

##### 3.1. DESIGN

This study is a longitudinal cohort study with a single proposed evaluation and the potential for further follow-up evaluations over five years. The study population will include people with HIV and age-matched people without HIV who participated in the APPLES study previously or are newly-recruited patients attending APPLES clinical sites or sites outside the APPLES network. Potential subjects will be approached for participation, consented and asked to participate in the cohort. As an initial proposed evaluation, participants will be asked to provide a single blood sample for testing for mutations associated with CH and to complete a survey regarding recent medical diagnoses and ART use. We will also ask that participants agree to follow-up research projects during the five year cohort study, which may involve more data collection including clinical samples.

We expect that 80% of our study participants will be have participated previously in APPLES, and about 20% will be newly-recruited.

##### 3.2. STUDY GROUPS

In this study there will be one group of 220 individuals with HIV, and one group of age-matched individuals without HIV.

##### 3.3. NUMBER OF PARTICIPANTS

The total number of expected participants in this study is 440.

##### 3.4. NUMBER OF SITES

This is a multi-centre interstate study. Currently we have six sites that have confirmed their intent to participate. We are continuing to approach others and anticipate that 10 sites total will participate. These sites were the highest-enrolling sites that participated in the APPLES study. The expected number of participants that will be enrolled in this study at each is the same as was enrolled into the previous APPLES study (440 total, 220 with HIV and 220 without HIV).

##### 3.5. DURATION

The total duration of the longitudinal cohort study is 5 years. After the initial proposed study visit to evaluate for clonal haematopoiesis, the first follow-up visit will occur 6-12 months later. Subsequently, study visits will be scheduled up to every 1-2 years (maximum total number of six study visits over five year duration). Subsequent visits will be determined by the results of this initial evaluation, and by developing research interests in the field of aging and HIV.

#### 4. PARTICIPANT SECTION

##### 4.1. INCLUSION CRITERIA

- Men and women aged >55
- For participants without HIV: an HIV negative test within 12 months prior to enrolment. If no HIV-negative test result is available within 12 months prior to enrolment, then participants will be tested for HIV as part of standard of care, if indicated by the guidelines for HIV testing published by the Australasian Society for HIV, Viral Hepatitis and Sexual Health Medicine[14]. This study will not conduct HIV testing; therefore, any participants without HIV being considered for participation in the study will need to have

had a standard of care HIV negative test within the past 12 months. Some subjects at on-going risk for HIV are recommended to have periodic HIV testing and may be due for such testing as part of standard of care, at the time of enrolment into the study.

- Willing and able to provide written informed consent and willingness to participate in and comply with a longitudinal cohort study including 1) consent to providing blood samples for full blood count, inflammatory marker testing and genomics analysis 2) consent to linking their data to national and state-wide data registries (including consent to providing personally identifying information); and 3) consent to participate in future follow-up studies

##### 4.2. Exclusion Criteria

- Unwilling or unable to provide consent to participate

#### 5. STUDY OUTLINE

##### 5.1. Investigation plan

The initial evaluation of the longitudinal cohort study to assess for clonal haematopoiesis will consist of one study visit where consent, enrolment and completion of study assessments will occur. At the enrolment visit, participants will give a blood sample and fill out a questionnaire, and sites will document information from the clinical record.

Chart Review

Sites will review the participant's chart and abstract information on the patients including: date of HIV diagnosis, medications the patient is taking (for example, anti-platelets, anti-hypertensive agents, lipid lowering agents, anti-diabetes agents). The most recent vital signs obtained at their last clinic visit will be collected on all participants. We will also ask sites to document the patient's medical diagnoses and any documentation of clinical events such as heart attack or stroke. Some laboratory data will also be documented including CD4 count and viral load (most recent) for participants with HIV. In addition, we will ask about ART history (for participants with HIV) and PrEP use (for participants without HIV) including when treatment was started and what medications the patient has been taking.

Survey

Participants will be asked to fill out a questionnaire on the following topics: use of tobacco, alcohol and recreational drugs; their medical history including any diagnoses and any clinical events (for example, myocardial infarction, cerebrovascular accident, or injuries); family history of medical problems/diagnoses; medications they are taking (to indicate from a list). For participants with HIV we will also ask about ART regimens and when they have been treated, to the best of their recollection. For example, we will ask them to indicate how long after their diagnosis they began HIV medication, and whether they have taken it consistently or had any major breaks in treatment since diagnosis. A specific question regarding use of stavudine or zidovudine will also be asked as these drugs are known to cause bone marrow dysplasia. We will also ask participants without HIV about use of pre-exposure prophylaxis (PrEP) to prevent HIV acquisition.

The questionnaire will be made available online or can be filled out on paper if the participant does not have access to the internet (the paper survey can be completed at their enrolment visit). For participants completing the survey online at home, they will be given a participant code (their unique study ID) and the URL link to the survey. Confidentiality of participants who join the study will be protected by the automatic survey system. Participant email addresses and/or phone numbers will be stored in a secure registration module separate from the questionnaires module.

Blood samples collection

For all participants, blood samples will be collected at enrolment to test for the following: the presence of mutations associated with CH, full blood count and specific inflammatory markers (D-dimer, hsCRP, cystatin C, and IL-6 and ESR). Once drawn at the study visit, blood samples will be sent by courier from the sites to an accredited pathology laboratory where they will be processed and FBC and inflammatory marker testing will be completed. Subsequently, whole blood and plasma samples will be sent to Peter MacCallum Cancer Centre in Melbourne for testing for somatic mutations associated with clonal haematopoiesis.

Sequencing methodology

We will use targeted amplicon deep sequencing (TS) using an amplicon panel designed across 55 known genes recurrently mutated in clonal haematopoiesis of indeterminate potential (CHIP), myelodysplastic syndromes (MDS) and acute myeloid leukaemia (AML) (Table 2). Further orthogonal validation of mutations will be performed using droplet digital PCR (ddPCR) using custom designed probes.

Table 2.

| **GENE** | | | | | | |
|---|---|---|---|---|---|---|
| ASXL1 | DIS3 | GNAS | MLL2 | PHF6 | SF3B1 | TP53 |
| BCOR | DNMT3A | IDH1 | MPL | PRPF40B | SMC1a | U2AF1 |
| BRAF | DOCK4 | IDH2 | MYC | PTEN | SMC3 | UMODL1 |
| CALR | ETV6 | JAK2 | NF1 | PTPN11 | SRSF2 | WAC |
| CBL | EZH2 | KDM6A | NPM1 | RAD21 | STAG2 | WT1 |
| CDKN2A | FLT3 | KIT | NRAS | RUNX1 | STAT3 | ZRSR2 |
| CSF1R | GATA1 | KRAS | PDGFRA | SETBP1 | SUZ12 | PPM1D |
| CUX1 | GATA2 | MLL | PDS5B | SF3A1 | TET2 | |

*Targeted Deep Sequencing*. DNA derived from plasma will be first subjected to pre-amplification[15]. DNA from whole blood will not be pre-amplified. Target specific amplification will then be performed using the Fluidigm Access Array™ system. Following amplification, products will be harvested, tagged with sample-specific barcodes, pooled together and purified using AMPure XP beads. All samples will be analysed in duplicate to control for PCR artefacts. The purified libraries were then sequenced on the Illumina MiSeq or NextSeq platform. Sequenced reads were mapped to the human reference genome (version hg19) using BWA-MEM (version 0.7.12) with default parameters. Variant calling and curation will be performed using an in-house pipeline (PATH-OS)[16]. Variants in the curated list will then be annotated based on their prognostic or functional relevance.

*Digital PCR*. Digital PCR will be performed using the Bio-Rad QX200 Droplet digital PCR system following manufacturer's protocols. Allele-specific PCR assays to specifically detect and quantify the fractional abundance of point mutations and corresponding wild-type alleles will be commercially obtained (PrimePCR™ PCR Primers and Assays, BioRad Laboratories). Data analysis will be carried out using the QuantaSoft Software, version 1.7 (Bio-Rad).

Future data collection and study visits. Depending on the results of our initial analysis, we may want to collect further information on participants such as more detailed laboratory information such as CD4 count and viral load, and more detailed clinical information such as ART history, concomitant medications, medical procedures, and hospitalizations. Therefore, we will schedule one follow-up visit with participants 6-12 months after the initial enrolment visit, and subsequent follow up visits every 1-2 years after that (total number of six study visits over five year duration). These visits may include clinical sample collection, further interview with the participant, non-invasive clinical testing (such as neurocognitive or exercise testing) or more detailed medical record review by the site. If any further procedures are planned, a protocol amendment will be submitted for HREC review and a new consent will be obtained from the participants. We may also obtain data through linkage to state and national databases. If data linkage studies are pursued, the appropriate ethics submissions will be completed with the appropriate state or national ethics committees including Population Health HREC, as appropriate.

These projects may require further funding and will depend on the results of the initial evaluation. The potential for future data collection will be described clearly in the PICF for the participants to review prior to providing consent to participate in the study.

##### 5.2. Study Procedure Risks

Samples acquired for this project will be used to evaluate the frequency and clinical impact of genetic mutations and epigenetic changes within the blood. It unlikely but possible that we may uncover a genetic mutation with implications for a subject’s family. In the panel of 55 genes that we are testing for (Table 2), there are three that are associated with known heritable cancer syndromes. These mutations are very rare in the general population and not likely to be detected in our study population. However, if a heritable genetic mutation is discovered, we will follow the guidelines set by NHMRC. The subject and the treating doctor will be informed that an inherited abnormality has been discovered and that they will need to have the results confirmed by standard-of-care genetic testing. They will be offered appropriate genetic counselling through the Familial Cancer Service at the Royal Melbourne Hospital or the Clinical Genomics Unit at St. Vincent’s Hospital in Sydney (both are external to the study team). In addition, we may recommend that the subject inform his/her relatives of the discovery, and may facilitate access to external genetic counselling, as required.

##### 5.3. Recruitment and Screening

Recruitment of participants with and without HIV will be through the existing network of clinical sites that participated in the APPLES study or through new sites outside the APPLES network. 440 participants (220 with HIV and 220 without HIV) will be included in this study including both subjects that participated previously in APPLES and subjects that are newly-recruited. Previously-recruited patients will be contacted via their preferred mode of contact (as stated on the initial APPLES PICF), or they will be approached as they present for routine care and asked if they would be interested in participating. Newly-recruited participants will be approached for participation consecutively by their providers, as they present for routine care and are identified as eligible for participation.

##### 5.4. Informed Consent Process

Privacy and confidentiality are key considerations for this study. Written informed consent will be obtained for all participants and a copy of the participant information and consent form (PICF) will be provided to each participant. Study personnel will carefully explain that consent is voluntary (free of coercion and pressure) and will review the study requirements with the participant. Participants will be given the opportunity to ask any questions of the principal investigator or other study personnel. And if they are happy to proceed, they will be asked to sign the PICF. We will seek informed consent specifically to record clinic data and questionnaire data as well as consent to record personal identifiers for data linkage and obtain blood samples for testing. Specific consent will also be obtained regarding collection of samples for genomics testing. Considerable efforts will be made to adequately inform participants of the importance of this study and the need to use full name identifiers, and of the procedures we will follow to maintain confidentiality.

As described in section 5.1., we will also ask participants to consent to possible participation in follow-up study visits that may be conducted. These study visits may include clinical record review, participant questionnaire completion, and collection of blood samples. This future research will depend on the initial findings of the first evaluation, and therefore may be done in a new area of research, or as follow-up studies to address questions related to clonal haematopoiesis. It is anticipated that patients may be contacted 6 months after the initial evaluation for the first follow-up visit, and subsequently every 1-2 years over the total duration of the longitudinal cohort study (5 years), for a total of 6 study visits. If the study evolves such that there are other evaluations proposed to be done outside of this time frame or outside of the above study procedures (blood sample collection, medical record review and questionnaire administration), we will submit a protocol amendment to the HREC as well as a new PICF and will re-consent the study participants to do these evaluations. Participants will be contacted by phone, SMS, email or regular mail as directed by the individual participant.

##### 5.5. Enrolment Procedure

The participant will be enrolled into the study after the informed consent process has been completed and the participant has met all inclusion criteria and none of the exclusion criteria. The participant will receive a study enrolment number and this will be documented in the participant's medical record and on all study documents.

#### 6. TISSUE COLLECTION/BIOBANKING

There is one proposed collection of blood in this study, with the possible for future collections at study follow-up visits, if scheduled (see above). Specific consent will be obtained from participants regarding the collection, storage and analysis of their blood samples for research purposes. There will be no samples obtained for this study to complete standard of care testing.

Sites will label samples with a patient name code and their unique study ID number. They will not include full name or any other personal identifiers. Samples will be transferred to an accredited pathology laboratory for testing for inflammatory markers and full blood count. The accredited pathology laboratory will also process the samples to plasma which will be subsequently sent to Peter MacCallum Cancer Centre for sequencing to identify mutations associated with clonal haematopoiesis. Samples sent to Peter MacCallum Cancer Centre will not be labelled with the name code, but only with the unique study ID and will therefore not contain any personally identifying information. Therefore, tissue samples will be individually re-identifiable but the privacy and confidentiality of the patients will be maintained when transferring samples to Peter MacCallum Cancer Centre. Further details on confidentiality are described in Section 10.

After testing, any remaining clinical sample material will be stored for seven years after the completion of the study analysis, for the purpose of further assessments within this longitudinal cohort (i.e. for this research into clonal haematopoiesis only). Samples will only be sent to an accredited pathology laboratory, St. Vincent's Centre for Applied Medical Research, or to Peter MacCallum Cancer Centre. If the samples collected for this study are to be used for another study, or are to be sent to another site, a protocol amendment will be submitted to the HREC and a new consent will be obtained from the participants.

#### 7. SAFETY

As this study only involves obtaining one blood sample, we do not anticipate any adverse events related to study procedures. As described above in Section 5.2, it is possible (though unlikely) that during the course of the project a genetic mutation may be identified that has implications for a subject's family. Please see Section 5.2 for further details.

##### 7.1. Early Termination

In the unlikely event that there is early termination of the study, Kirby Institute will be responsible for coordinating with the sites to ensure that they contact and inform the participants. The Kirby will immediately notify the HREC and will compile a final study report, as per standard practice.

#### 8. OUTCOMES AND FUTURE PLANS

As the results of genetic testing to identify somatic mutations are for research purposes only and are not standard of care clinical practice, the results of this study will not be returned to the participants.

Study governance. The study will be coordinated by the Kirby Institute and there will be a single Protocol Steering Committee (PSC). The PSC will include a representative from each participating site and other representatives at the Kirby including a biostatistician. The PSC will seek the expertise of other key opinion leaders as necessary. The PSC will be the primary management entity for the collaborative study group. This group will meet by teleconference, arranged as required and at least twice per year during the study. Decisions in this group will be reached by consensus among the designated membership. Routinely the views of other stakeholders will be sought at meetings of the PSC and this will allow others to attend meetings. The PSC, comprised of members from a range of internal and external stakeholders will guide the design, implementation and conduct of the study and consider requests from external investigators for access to study bio-specimens.

Publication policy. The primary manuscript publishing the results of our first analysis of clonal haematopoiesis will be from the "ARCHIVE Study Group" which will include a select group of members of the PSC (based on interest and accrual) and specified external collaborators who will liaise with the study team. All PSC members and other key personnel at study sites that are not listed as named authors will be acknowledged in the manuscript.

There are currently no anticipated secondary uses of the data. Potential future use of data may include other studies to evaluate HIV and aging and/or the mechanisms behind the accelerated development of comorbidities and clinical events in persons with HIV.

#### 9. STATISTICAL CONSIDERATIONS

Sample size calculation. We will recruit 440 participants: 220 with HIV and 220 without HIV. We will assume that 10% of our participants without HIV will have somatic mutations associated with clonal haematopoiesis, which is consistent with published studies in the literature reporting a range of 6-10%[12,13,17]. Emerging data suggest that with the increasing sequencing depth that will be used in this study, detection of mutations is greatly increased from that reported in the seminal papers above. Therefore, we will assume we will detect mutations among 10% of participants without HIV and will power our study to detect a clinically interesting increase in prevalence (doubling) from 10% to 20% prevalence among participants with HIV. Testing 220 participants without HIV and 220 participants with HIV will give us 80% power to detect an increase to 20% prevalence of mutations among participants with HIV compared to the background mutation rate of 10% among participants without HIV.

Statistical analysis plan. The prevalence of somatic mutations associated with clonal haematopoiesis will be summarized by HIV infection status. Conventional statistics will be used to compare the results across the two groups and determine if there are any statistically significant differences.

We will also test all participants' plasma samples for the presence of these mutations and compare it to those found in the whole blood testing. This analysis will be done using a correlation analysis comparing mutant allele fraction of whole blood with plasma using the Spearman method[18]. The purpose of this comparison is to validate using plasma samples for detection of somatic mutations. Our evaluation of an association between somatic mutations and inflammatory markers and clinical outcomes (comorbidities and medical events) will be exploratory.

Health information collected for this study may also be linked to other state and national databases such as; the Admitted Patients Data Collection, Registry of Births, Death and Marriages, and the NSW Cancer Registry. This linkage will depend on the requirements of this and future research projects.

#### 10.CONFIDENTIALITY AND STORAGE AND ARCHIVING OF STUDY DOCUMENTS

Participant confidentiality will be maintained by recording data in two different data sets. The first will be created from the original APPLES dataset that includes full identifiers of participants along with a Unique identifier (UNID) for the study. The second dataset with all research data for this study will only include the patient UNID. All patient information will be stored on secure, password access only, electronic databases on the server at the Kirby Institute. Access to the complete patient identifiers will be limited to the Principal Investigators and data management administrators of the study. This file will only be used in the future for data linkage or in the case of specific further testing based on the results of the initial study and will follow appropriate ethical procedures as required for linkage.

Laboratory samples will be stored only with the assigned UNID. Samples will be stored at the accredited pathology laboratory and at Peter MacCallum Cancer Centre in a secure facility …. All data generated by the study will be kept for 7 years, and after that for as long as the data are considered to have scientific utility. Participant data will only be used for the purpose of this study.

#### 11. OTHER STUDY DOCUMENTS

Documents included with this submission are:

- Questionnaire for study participants (this questionnaire is non-validated, but based on the 45 and up study[19], which is a well-known study in NSW)

#### 12. RESOURCES

This study is funded by the Kirby Institute for Infection and Immunity in Society at the UNSW Sydney, Australia (Kirby). Kirby will supply funds for the following: 1) site costs associated with enrolment of patients, completion of study assessments, and shipment of samples; 2) costs associated with testing samples for inflammatory markers, storage of samples at St. Vincent's Centre for Applied Medical Research (AMR) in Sydney, New South Wales or VIDRL in Melbourne, Victoria; and 3) shipment of samples from AMR and VIDRL to Peter MacCallum Cancer Centre in Melbourne, Victoria. Peter MacCallum Cancer Centre will supply funds for testing all samples for mutations associated with clonal haematopoiesis.

#### 13. REFERENCES

1. Vance DE, Mugavero M, Willig J, Raper JL, Saag MS. Aging with HIV: a cross-sectional study of comorbidity prevalence and clinical characteristics across decades of life. J Assoc Nurses AIDS Care 2011;22:17-25.

2. Schouten J, Wit FW, Stolte IG, et al. Cross-sectional comparison of the prevalence of age-associated comorbidities and their risk factors between HIV-infected and uninfected individuals: the AGEhIV cohort study. Clin Infect Dis 2014;59:1787-97.

3. Hasse B, Ledergerber B, Furrer H, et al. Morbidity and aging in HIV-infected persons: the Swiss HIV cohort study. Clin Infect Dis 2011;53:1130-9.

4. Guaraldi G, Orlando G, Zona S, et al. Premature age-related comorbidities among HIV-infected persons compared with the general population. Clin Infect Dis 2011;53:1120-6.

5. Wada N, Jacobson LP, Cohen M, French A, Phair J, Munoz A. Cause-specific life expectancies after 35 years of age for human immunodeficiency syndrome-infected and human immunodeficiency syndrome-negative individuals followed simultaneously in long-term cohort studies, 1984-2008. Am J Epidemiol 2013;177:116-25.

6. Deeks SG. Immune dysfunction, inflammation, and accelerated aging in patients on antiretroviral therapy. Top HIV Med 2009;17:118-23.

7. Dolcetti R, Giagulli C, He W, et al. Role of HIV-1 matrix protein p17 variants in lymphoma pathogenesis. Proc Natl Acad Sci U S A 2015;112:14331-6.

8. Friis-Moller N, Weber R, Reiss P, et al. Cardiovascular disease risk factors in HIV patients--association with antiretroviral therapy. Results from the DAD study. AIDS 2003;17:1179-93.

9. Lange RA, Hillis LD. Cardiovascular complications of cocaine use. N Engl J Med 2001;345:351-8.

10. Prestage G, Jin F, Kippax S, Zablotska I, Imrie J, Grulich A. Use of illicit drugs and erectile dysfunction medications and subsequent HIV infection among gay men in Sydney, Australia. J Sex Med 2009;6:2311-20.

11. Antiretroviral Therapy Cohort C. Life expectancy of individuals on combination antiretroviral therapy in high-income countries: a collaborative analysis of 14 cohort studies. Lancet 2008;372:293-9.

12. Genovese G, Kahler AK, Handsaker RE, et al. Clonal hematopoiesis and blood-cancer risk inferred from blood DNA sequence. N Engl J Med 2014;371:2477-87.

13. Jaiswal S, Fontanillas P, Flannick J, et al. Age-related clonal hematopoiesis associated with adverse outcomes. N Engl J Med 2014;371:2488-98.

14. Australasian Society for HIV, Viral Hepatitis and Sexual Health Medicine National HIV Testing Policy. Indications for HIV testing. at http://www.testingportal.ashm.org.au/hiv/indications-for-hiv-testing.)

15. Dawson S-J, Tsui DWY, Murtaza M, et al. Analysis of Circulating Tumor DNA to Monitor Metastatic Breast Cancer. New England Journal of Medicine 2013;368:1199-209.

16. Wong SQ, Waldeck K, Vergara IA, et al. UV-Associated Mutations Underlie the Etiology of MCV-Negative Merkel Cell Carcinomas. Cancer Res 2015;75:5228-34.

17. Xie M, Lu C, Wang J, et al. Age-related mutations associated with clonal hematopoietic expansion and malignancies. Nat Med 2014;20:1472-8.

18. Yeh P, Dickinson M, Ftouni S, et al. Molecular disease monitoring using circulating tumor DNA in myelodysplastic syndromes. Blood 2017;129:1685-90.

19. Sax Institute. 45 and Up Study. . Sax Institute (Accessed May 26 2017, at https://www.saxinstitute.org.au/our-work/45-up-study/.)

NHMRC National statement on ethical conduct in human research (2007) – updated May 2015, available at: https://www.nhmrc.gov.au/guidelines-publications/e72

NHMRC Australian code for the responsible conduct of research, available at https://www.nhmrc.gov.au/guidelines-publications/r39
